# Oestradiol, emotion regulation and the limbic system: effects on grey matter volume

**DOI:** 10.1101/2025.09.16.25335855

**Authors:** A.F. Denninger, E. Rehbein, I. Sundström-Poromaa, B. Derntl, L. Kogler

## Abstract

Mastering emotion regulation is crucial for social skills and mental health. Hormonal fluctuations, particularly in oestradiol levels (E2) across the menstrual cycle, significantly impact emotion processing. E2 is known to influence emotion regulation, mental health, and the plasticity of limbic and striatal regions, which are involved in emotion processing and are rich in E2 receptors. Although, research indicate that E2 levels may impact grey matter volume (GMV) of limbic and striatal areas, sufficient causal evidence is missing so far. Further, because of the additional fluctuations of progesterone across the menstrual cycle, the sole impact of E2 on brain volume has been difficult to disentangle.

To isolate the effects of E2 from other fluctuating sex hormones, we employed a randomised placebo-controlled, double-blind, cross-over design and administered oral E2 to 27 naturally cycling females during their early follicular phase (low endogenous sex hormones levels). We analysed emotion regulation strategies and E2 levels to assess their impact on regional grey matter volume (GMV).

Our data showed that a rapid increase of E2 is negatively associated with bilateral striatal GMV. Moreover, greater use of reappraisal is associated with reduced GMV of the striatum. Rapidly increased E2 did not influence GMV of other limbic regions.

These results highlight that rapid increases in E2 and individual differences in emotion regulation dynamically modulate limbic GMV. This offers important implications for female mental and brain health during associated with hormonal fluctuations. Considering female endocrine profiles improves to a hormone-informed health care and therefore supports individualized medicine.

## 1. Introduction

Oestradiol (E2) is traditionally viewed as a reproductive hormone, synthesized primarily in the ovaries and, to a lesser extent, in adipose tissue of females (Barakat et al., 2016). Importantly, E2 is also synthesized de novo within the brain (Azcoitia et al., 2011), where peripheral and neuro-derived E2 can act locally via estrogen receptors (ERs) to exert neuromodulatory (Bayer et al., 2020; Rehbein et al., 2021) and potential neuroprotective effects (Arevalo et al., 2015), driving morphological and neurochemical changes. Oestradiol-induced effects in the brain include modulation of synaptic plasticity, regulation of neurotransmitter systems, reduction of oxidative stress and inflammation, promotion of neuronal growth, plasticity via regulation of brain-derived neurotrophic factor (BDNF), cell signalling pathways, and transcription factor activation (Arevalo et al., 2015).

However, E2 is a hormone in constant change throughout the female reproductive life span. Endogenous E2 fluctuations as well as phases of altered E2 exposure modulate neuronal emotion regulation, emotion processing and functional connectivity of the emotion regulation network (Derntl et al., 2024; Rehbein et al., 2021; Sundström-Poromaa, 2018). During the menstrual cycle, E2 levels are low in the early follicular phase, peak shortly before ovulation, and then vary together with progesterone (P4) during the luteal phase, before both hormones rapidly decline just before menstruation (Roos et al., 2015). Individuals can regulate their emotions by applying a variety of strategies and studies indicate that the use of emotion regulation strategies varies in effectiveness between menstrual cycle phases. Reappraisal, a cognitive emotion regulation strategy, is based on changing the meaning of situations to alter the emotional responsiveness (Gross, 2015). Reappraisal successfully reduces negative affect (Buhle et al., 2014) and higher E2 levels are associated with more effective reappraisal use (Graham et al., 2017). Consequently, when E2 levels are low (as in the early follicular phase), individuals show increased effort to apply reappraisal but with reduced success (Wu et al., 2014). Rumination on the other hand is a regulation strategy with a repetitive and recurrent focus on negative thoughts or emotions (Smith & Alloy, 2009), and is often associated with increased anxiety and stress levels (Aldao et al., 2014; Harrington & Blankenship, 2002), and consequently, poor mental health (Graham et al., 2018; Nolen-Hoeksema, 2000). Research has shown that females with low E2 who engage in rumination experience greater negative affect (Graham et al., 2018). Some evidence also suggests that emotion regulation traits such as reappraisal influence feelings of sadness differently throughout the menstrual cycle (Wu et al., 2019). Similarly, Rafiee et al. (2023) demonstrated a negative association between emotion regulation traits and emotion recognition when progesterone levels were elevated. These findings suggest that individual differences in emotion regulation abilities may shape emotional experiences in the context of hormonal changes.

E2 administration leads to alterations in activity patterns during reappraisal use (Rehbein et al., 2021) and changes in functional network connectivity (Derntl et al., 2024). On a neuronal level, emotion processing and regulation engage key brain regions, particularly limbic structures such as the hippocampus, amygdala, anterior cingulate cortex (ACC), and regions closely connected to them such as the striatum (Etkin et al., 2015; Morawetz et al., 2017; Wu et al., 2019). These areas are central to affective processing, motivation, and the regulation of behavioural and physiological responses to emotional stimuli (Etkin et al., 2015; Morawetz et al., 2017). Importantly, these regions are also rich in ERs, indicating that they may be sensitive to changes in E2 levels (Almey et al., 2015; Krentzel et al., 2021). This E2 fluctuation-specific responsivity may involve changes in synaptic structure and function (Azcoitia et al., 2018), and therefore might be associated with affect and cognition (Azcoitia et al., 2018; Mosconi et al., 2024). Changes in grey matter volume (GMV) have been observed in relation to habitual use of emotion regulation strategies. Reappraisal has been negatively associated with ventral striatum GMV (Yoon & Jung, 2025) and positively associated with amygdala GMV (Hermann et al., 2014). Regarding rumination, positive associations have been reported with GMV in the para-hippocampus (Wang et al., 2015) and the caudate nucleus (part of the striatum; Joss et al., 2025), while negative associations emerged with the left ACC GMV (Kühn et al., 2012). It should be noted that the proportion of female participants varied in these studies, as did their reproductive and hormonal status (from adolescents to postmenopausal women), which may have influenced the observed associations between emotion regulation traits and GMV.

Across the menstrual cycle, high E2 phases (like the pre-ovulatory phase) are linked to increased GMV in the hippocampus (Lisofsky et al., 2015; Zsido et al., 2023) and ACC (Protopopescu et al., 2008), but decreased GMV in the striatum (Protopescu et al., 2008; Pletzer et al., 2018) in comparison with low E2 phases (like the early follicular or late luteal phase). Findings for the amygdala are, however, mixed (Meeker et al., 2020; Ossewaarde et al., 2013; Zwaan et al., 2022). Taken together, these findings suggest that hippocampus, ACC, amygdala, and the striatum exhibit structural plasticity in response to endogenous fluctuating E2 levels, with higher GMV of the hippocampus and ACC and lower GMV of the striatum when E2 levels are high vs. low.

Although an association between E2 levels and the application of emotion regulation strategies is assumed, the literature is scarce regarding how E2 and different emotion regulation traits affect GMV in emotion processing regions, and if these two variables interact in their influence on GMV of these regions. With the current study we therefore aim to investigate the effect of E2 and emotion regulation traits on GMV of regions involved in emotion processing. By experimentally increasing E2 levels in a randomized placebo-controlled cross-over design during the early follicular phase, we were able to compare elevated E2 levels to naturally low E2 levels and investigate the variability in E2 levels, which appear to be the driving factor for structural changes rather than absolute levels (Zwaan et al., 2022). Additionally, the assessment of reappraisal and rumination trait use gives us insight into the impact of E2 levels on brain plasticity in females applying reappraisal or rumination. With this design, we wanted to examine whether (a) increasing E2 levels are positively associated with greater GMV in the hippocampus and ACC (De Bondt et al., 2013; Lisofsky et al., 2015), and negatively associated with GMV in the amygdala and striatum (Ossewaarde et al., 2013; Pletzer et al., 2018; Protopopescu et al., 2008); and (b) whether the use of cognitive emotion regulation strategies such as reappraisal and rumination, differentially affects brain structure. Given the limited and inconclusive literature, we exploratorily hypothesized that amygdala, hippocampus, ACC and striatum GMVs would be associated with emotion regulation strategies. Specifically, we hypothesised that trait use of cognitive reappraisal would be positively associated with larger amygdala volume (Hermann et al., 2014), and negatively with smaller striatum volumes (Yoon & Jung, 2025). Rumination would be associated with larger striatum (Joss et al., 2025) and smaller ACC GMV (Kühn et al., 2012). Additionally, we hypothesized that the trait use of emotion regulation strategies would interact with E2 levels influencing brain volume. For cognitive reappraisal we assumed that higher trait use would be associated with smaller GMV of the striatum when E2 levels are high. For the other regions no direct hypothesis could be formulated.

## 2. Experimental procedures

### 2.1. Sample description

Inclusion criteria were female sex assigned at birth, age 19-35 years, right handedness and a regular menstrual cycle lasting between 26-32 days. To ensure the regularity of the menstrual cycle, two cycles were tracked before participation. Participants were excluded if they had any MRI-contraindication (e.g., metal implants), present or past mental (assessed via the SCID, Wittchen et al., 1997), neurological or endocrine disorders, use of hormonal contraceptives in the last six months, medication intake, past or present pregnancies. All participants signed an informed consent from. The study was approved by the Ethics Committee of the Medical Faculty, University of Tübingen (754/2017/BO1), preregistered NCT06312033) and data was collected in Tübingen, Germany, between 2018 and 2019 as part of a larger scope.

### 2.2. Procedure

We applied a randomised placebo-controlled, double-blind, cross-over design for the E2 administration. After an initial screening and assessment of variables for sample description (verbal intelligence (WST, Schmidt & Metzler, 1992), speed processing and cognitive flexibility (TMT-A/B, Rodewald et al., 2012), emotion regulation (HFERST, Izadpanah et al., 2019), participants reported the onset of their menstruation and were invited to the study within 2 to 5 days after menstruation onset to capture the early follicular phase. To safely and efficiently increase E2 levels comparable to pre-ovulatory levels (Bayer et al., 2020), a first dose of E2-valerate (6 mg, Progynova21©, Bayer Weimar Gmbh und Co.KG) or placebo pills (PLAC) were administered approximately 24h before the magnetic resonance imaging (MRI) assessment in a randomized double-blind manner. A second dose was administered approximately 6h before the MRI assessment to achieve peaking E2 levels during neuroimaging (Ndefo & Mosely, 2010). Before the MRI assessment, depression symptoms (BDI-2, Hautzinger et al., 2006) and state as well as trait anxiety (STAI, Laux et al., 1981) were assessed. After a wash-out period of at least two months the participants crossed over to the other drug condition (E2/PLAC) and the procedures were repeated (Fig. 1). As this study was part of a larger study, please also see Rehbein et al. (2021; 2022) and Derntl et al. (2024) for more detailed descriptions of the study procedure.

**Fig. 1.**
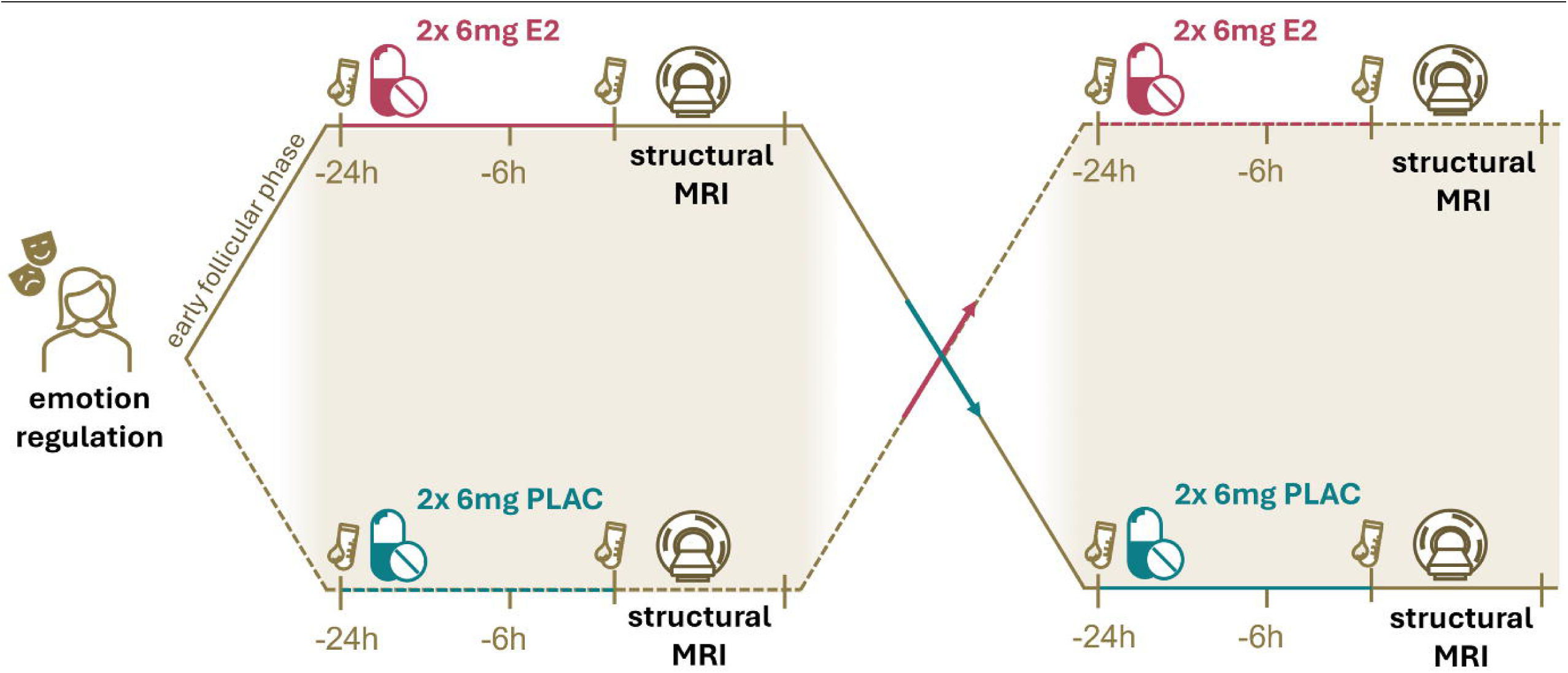
Study procedure. Assessment of trait use of emotion regulation including reappraisal and rumination followed by two consecutive drug administrations (E2 or PLAC) 24h and 6h before a structural MRI scan. Before drug administration and after MRI measurement blood samples were retrieved to assess hormone levels. This procedure was repeated after min. 2 months with the other drug condition.

### 2.3. Hormone assessment

Blood samples of hormone levels (E2, P4, testosterone) were collected before E2/PLAC intake and after the MRI session by the central laboratory, University Hospital Tübingen, and analysed by use of enzyme-linked immune-assay (ELISA). Sensitivity ranges were as follows, E2 = 43.6 -11,010 pmol/l; P4 = 0.67 – 190.8 nmol/l; testosterone = 0.24-52.05 nmol/l.

### 2.4. Neuroimaging

Structural MRI data was acquired by a 3T scanner (Prisma, Siemens, University Hospital Tübingen). T1-weighted images were obtained with a standard magnetization-prepared rapid gradient-echo sequence (TR = 2,300ms; TE = 4.16ms, slice thickness = 1.00mm, voxel size = 1x1x1mm, flip angle of 9°, distancing factor 50%, GRAPPA acceleration factor, sagittal orientation) and a 64-channel head coil.

Data was pre-processed applying the CAT12 toolbox (https://neuro-jena.github.io/cat/; SPM12 (https://www.fil.ion.ucl.ac.uk/spm/software/spm12; Matlab Mathworks, Natick, MA, USA). T1 weighted images underwent normalisation, longitudinal segmentation into cerebrospinal fluid, white matter volume and GMV, warping to MNI space, and smoothing with a full width at half maximum (FWHM) isotropic gaussian kernel of 8mm. Total intracranial volume (TIV) and GMV were extracted.

Region of interests (ROI) (Fig. 2) included bilateral hippocampus, ACC, amygdala and striatum. To inspire future studies, we further exploratorily investigated subregions (dorsal, ventral, and caudoventral striatum, (para-)hippocampus, dorsal, pre- and subgenual ACC) which are reported in the supplementary (S1). ROI masks were defined according to the JUBrain toolbox (Eickhoff et al., 2005), except for the striatum, which was defined according to Liu et al., (2020). Masks were further extracted with an adapted get_totals script (G. Ridgeway; http://www0.cs.ucl.ac.uk/staff/gridgway/vbm/get_totals.m) as done previously (Pletzer et al.,

**Fig. 2.**
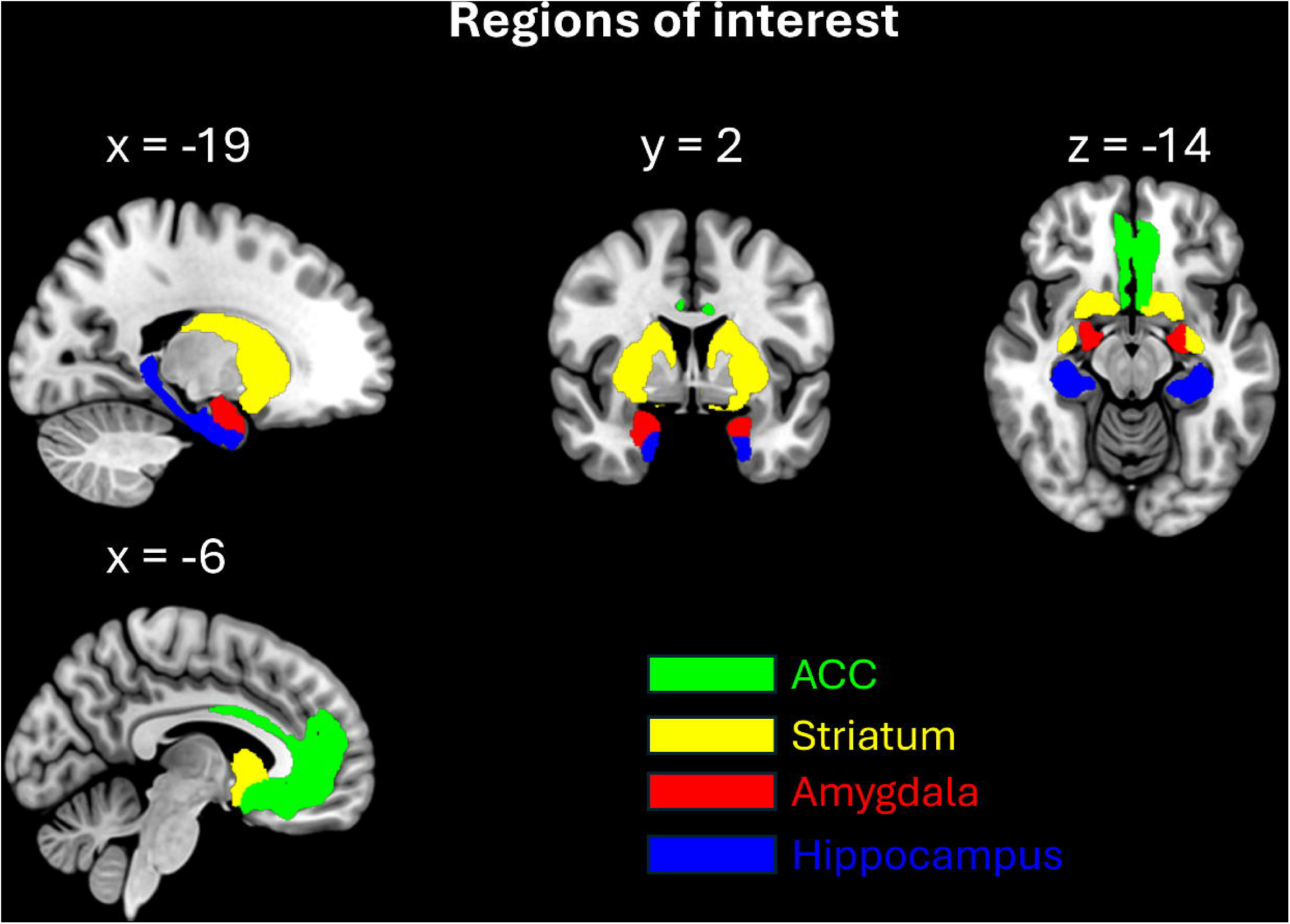
Region of interest. (ROI) in MNI-space. ACC (green), striatum (yellow), amygdala (red) & hippocampus (blue).

2018).

### 2.5. Statistical analysis

Statistical analyses were performed in Matlab (MathWorks R2024). Differences between drug conditions for behavioural and hormonal data were examined by dependent t-tests or Wilcoxon signed-rank test when assumptions for parametric testing were not met. Nonparametric effect size was calculated using Rosenthal’s r, where 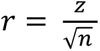, following Rosenthal (1991).

To assess the effect of a) E2 increase in the E2 or PLAC condition (E2 levels pre minus post drug administration, E2Δ) and b) emotion regulation trait (reappraisal, rumination) on GMV, individual robust multiple linear regression models were applied with MATLAB’s built-in robust model option (less sensitive to outliers). TIV and age were applied as covariates resulting in a) GMV ∼ E2Δ_E2 | PLAC_ + age + TIV; and b) GMV ∼ emotion regulation trait _reappraisal | rumination_ + age + TIV; and c) GMV ∼ E2Δ _E2 | PLAC_ * emotion regulation trait _reappraisal | rumination_ + age + TIV. E2 increase was expressed as positive values and log transformed. All predictors and covariates were centred relative to their mean. Multiple linear regression was selected based on the reduced risk of overfitting given our small sample size and the fact that we analysed the two drug conditions separately. To evaluate differences between the drug conditions beta values were calculated, Z-transformed and then tested for significance applying the cumulative distribution function of the standard normal distribution.

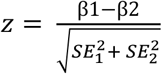

Level of significance was set to p = .05 and corrected for hemispheres applying false discovery rate (FDR) using the Benjamini and Hochberg (1995) procedure. Sample size was determined via G-power analysis reported by Rehbein et al. (2021).

## 3. Results

### 3.1. Sample description

Initially, 32 naturally cycling females participated in the study. Five participants had to be excluded due to missing MRI-data (n = 1), start of hormonal contraception use between the two sessions (n = 1) and depressive symptoms at one of the two drug-condition sessions (BDI-2 scores ≥ 20, n = 3). Thus, the final sample size included 27 females. Sample description including mean age, verbal and cognitive performance are reported in Tab. 1. State anxiety (t(26) = -.042, p = .97) and BDI-2 scores (t(26) = -.189, p = .85) prior to drug administration did not differ between drug conditions and effects did not depend on order of drug administration (Tab. 1).

**Tab. 1.** Sample description.

| | Mean ± SD | E2: Mean ± SD | PLAC: Mean ± SD | p- value |
| --- | --- | --- | --- | --- |
| **Age [years]** | 23.22 ± 3.0 | | | |
| **Verbal intelligence** | 32.59 ± 3.2 | | | |
| **Processing speed [s]** | 19.93 ± 4.1 | | | |
| **Cognitive flexibility [s]** | 36.94 ± 10.3 | | | |
| **Reappraisal trait** | 15.15 ± 2.7 | | | |
| **Rumination trait** | 14.07 ± 3.5 | | | |
| **Anxiety – trait** | 33.59 ± 7.4 | | | |
| **Anxiety – state** | | 38.04 ± 6.4 | 38.11 ± 8.1 | .967 |
| **Depression score** | | 6.26 ± 4.9 | 6.33 ± 5.3 | .852 |

Verbal intelligence (WST), processing speed (TMT-A), cognitive flexibility (TMT-B), reappraisal trait (HFERST), rumination trait (HFERST) and anxiety trait (STAI). Standard deviations (SD) are indicated.

### 3.2. Effective elevation of E2 levels

E2 levels were similar prior to E2/PLAC administration (Z = .17, p = .867, r = .03) and between pre and post PLAC drug administration (Z = -1.46, p = .145, r = -.29). Following E2 administration, E2 levels were effectively increased, both compared to baseline (Z = -4.02, p < .001, r = -.88) and to PLAC drug administration (Z = 3.82, p < .001, r = .88). Progesterone and testosterone levels remained low, see supplementary (Tab. S1, Fig. S2).

### 3.3. E2 increase is negatively associated with striatum volume

Multiple linear regression analyses revealed that in the E2 drug condition, E2Δ was negatively associated with bilateral striatal GMV (model_left_ R^2^ = .742, p < .001, β_left_ = -.685, p_left_ = .026; model_right_ R^2^ = .803, p < .001, β_right_ = -.854, p_right_ = .004). No significant association with striatal GMV was found during the PLAC drug condition (model_left_ R^2^ = .745, p < .001, β_left_ = .009, p_left_ = .908; model_right_ R^2^ = .729, p < .001, β_right_ = -.009, p_right_ = .921). The effect of E2Δ on bilateral striatum was significantly different between the E2 and PLAC conditions (Z_left_ = -2.676, p_left_ = .008; Z_left_ = - 3.374, p_right_ < .001), showing a stronger negative association during the E2 drug condition (Fig. 3, Tab. S3). No associations between E2 or PLAC drug administration was noted in the GMV of amygdala, hippocampus, and ACC. Parameters and median E2Δ are listed in the supplementary (Tab. S2 & 3).

**Fig. 3.**
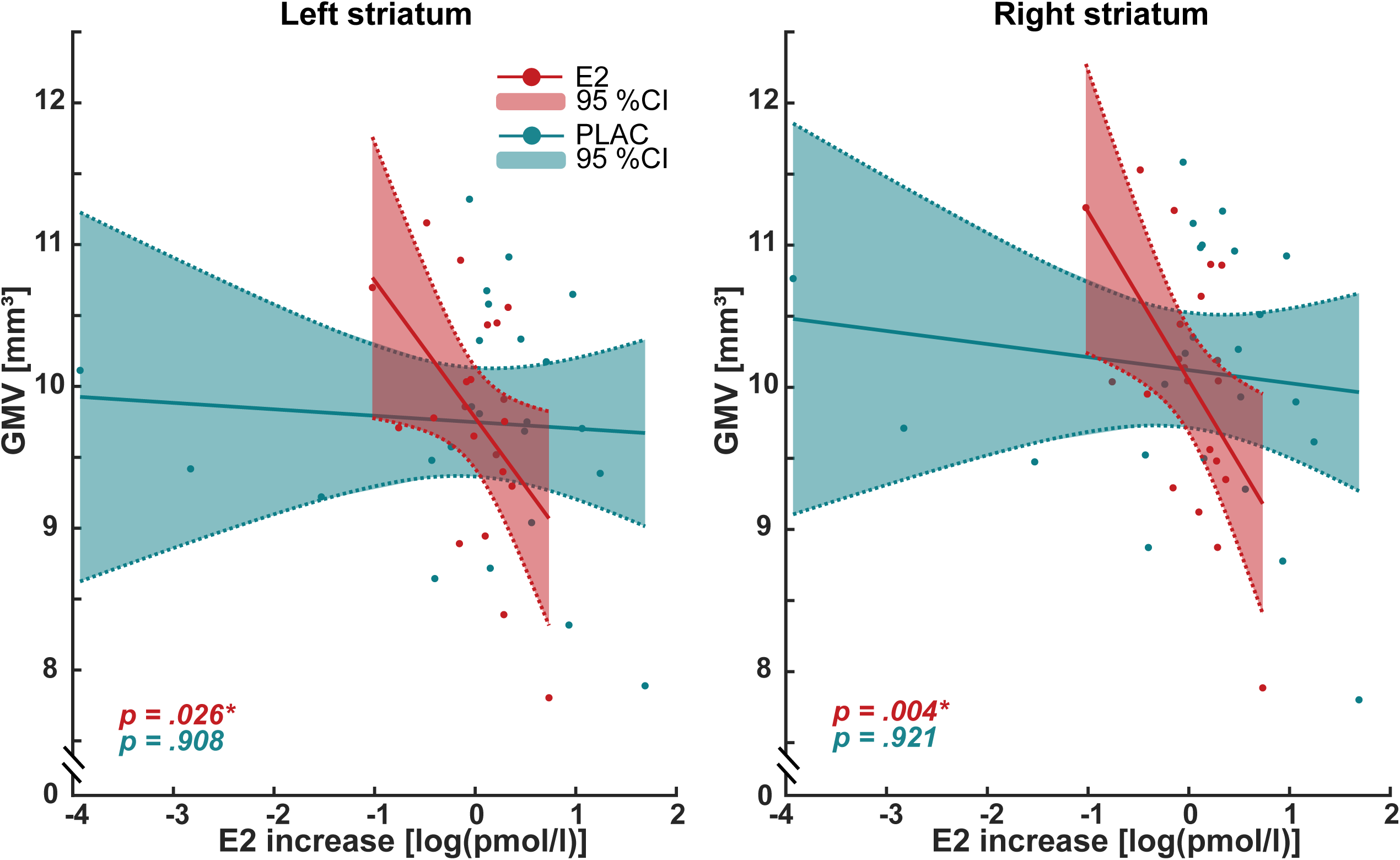
Association between log centred E2 increase (E2Δ) and bilateral striatum GMV [mm^3^] during E2 (red) and PLAC (blue) administration. 95% confidence intervals and significance levels are indicated. Note, that the E2 increase is centred in relation to its mean for each E2 and PLAC condition (0 = mean).

Exploratory analysis of striatal sub-regions showed a similar pattern for the right ventral (model_right_ R^2^ = .760, p < .001, β_right_ = -.110, p_right_ = .013), right caudoventral (model_right_ R^2^ = .567, p < .001, β_right_ = -.044, p_right_ = .023) and bilateral dorsal striatum (model_left_ R^2^ = .669, p < .001, β_left_ = -.584, p_left_ = .029; model_right_ R^2^ = .778, p < .001, β_right_ = -.073, p_right_ = .006) as for the whole striatum during the E2 drug condition (Tab. S3).

### 3.4. Emotion regulation trait affects striatal GMV, independent of drug condition

#### Reappraisal

Trait reappraisal was negatively associated with GMV of the bilateral striatum in the E2 (model_left_ R^2^ = .653, p < .001, β = -.102, p_left_ = .032; model_right_ R^2^ = .689, p < .001, β = -.114, p_right_ = .012) and PLAC drug condition (model_left_ R^2^ = .681, p < .001, β = -.081, p_left_ = .049; model_right_ R^2^ = .689, p < .001, β = -.090, p_right_ = .041) (Fig. 4). Beta values did not differ significantly between the E2 and PLAC drug conditions for bilateral striatum GMV (Z_left_ = -0.390, p_left_ = .696; Z_left_ = -.454, p_right_ = .650), suggesting an equal relationship across conditions (Fig. 4, Tab. S4). Reappraisal was not significantly associated with GMV of the amygdala, hippocampus and ACC in neither drug condition (E2: p_amyL_ = .715; p_amyR_ = .715; p_accL_= .896; p_accR_= .921; p_hipL_= .944; p_hipR_= .757; PLAC: p_amyL_ = .337; p_amyR_ = .559; p_accL_= .479; p_accR_= .952; p_hipL_= .952; p_hipR_= .460). Paramteres are listed in the supplementary (Tab. S4).

**Fig 4.**
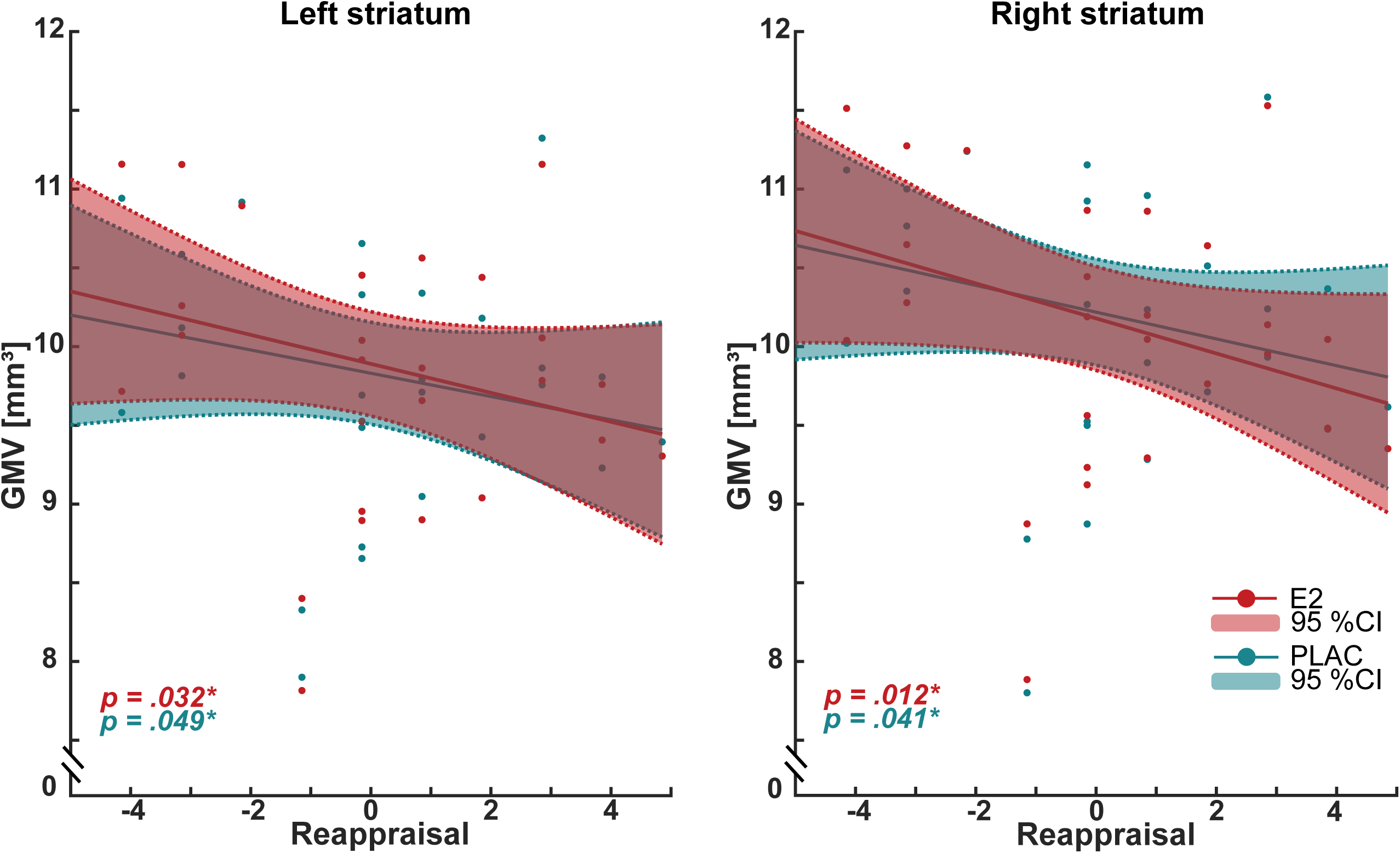
Association between emotion regulation strategy, reappraisal, and bilateral striatum GMV [mm³] during E2 (red) and PLAC (blue) administration. 95% confidence intervals and significance levels are indicated. Note, that reappraisal is centred relative to its mean (0 = mean).

Exploratory analyses on striatal sub-regions revealed that reappraisal was negatively associated with right ventral (model_right_ R^2^ = .628, p < .001, β_right_ = -.012, p_right_ = .050) and bilateral caudoventral (model_left_ R^2^ = .643, p < .001, β_left_ = -.005, p_left_ = .018; model_right_ R^2^ = .606, p < .001, β_right_ = -.006, p_right_ = .005) and dorsal (model_left_ R^2^ = .630, p < .001, β_left_ = -.092, p_left_ = .070; model_right_ R^2^ = .667, p < .001, β_right_ = -.085, p_right_ = .024) striatum GMV. However, slopes did not differ between the E2 and PLAC drug conditions (see supplementary Tab. S4).

#### Rumination

Trait rumination was not associated with GMV of any ROI (nor for any sub-regions) during either drug condition (p >.05). Parameters are listed in the supplementary (Tab. S5).

#### Interaction

No significant interactions between E2 and emotion regulation trait appeared (p > .05). Complete statistical parameters are found in the supplement (Tab. S6 & S7).

## 4. Discussion

In the current randomised placebo controlled cross-over study we aimed to identify the rapid effect of E2 on brain plasticity in naturally cycling females. Additionally, we wanted to explore the effect of E2 on brain anatomy in females applying reappraisal and rumination in everyday life. First, we demonstrated that the reliable and rapid increase of E2 levels was negatively associated with bilateral striatal GMV, with the relationship being more pronounced in the E2 drug than PLAC drug condition. Second, trait reappraisal use was negatively associated with striatal GMV; however, this effect was independent of E2 levels.

### 4.1. E2 affects striatal volumes

Our data shows that the rapid increase in E2, which mimics the pre-ovulatory surge in the middle of the menstrual cycle, impacts brain plasticity particularly in bilateral striatum: a greater increase in E2 was associated with a greater decrease in GMV. This negative association differed significantly from the PLAC drug condition. Thus, by using a placebo-controlled cross-over design during a phase of low progesterone and testosterone levels, our findings indicate that elevated E2 levels are causally associated with decreased GMVs of the striatum.

Indeed, our data aligns well with findings from the menstrual cycle, where hormone levels naturally fluctuate. The putamen (as part of the dorsal striatum) decreases in size pre-ovulatory, when E2 levels are high, compared to the luteal phase (Pletzer et al., 2018). Similar results were observed by Protopopescu et al. (2008) who showed decreased right putamen GMV during the late follicular phase (rising E2 levels). This is further supported by studies examining the effects of exogenous sex hormone administration via birth control pills — which typically administer a synthetic oestrogen such as ethinyl-oestradiol (combined with a synthetic progestin) for three weeks, followed by a one-week pill-free interval. During this pill-free interval, when hormone levels decline to levels comparable to the early follicular phase (Rodriguez et al., 2024), caudate GMV (as part of the ventral striatum) increase (De Bondt et al., 2013). Taken together with the current data, this suggests that the striatum and its subregions are sensitive to fluctuations in both exogenous and endogenous E2 variations.

Animal studies have shown that E2 can rapidly influence the dorsal striatum (Becker, 1990; Becker & Rudick, 1999) by modulating neuroplastic mechanisms including cellular proliferation (Wang et al., 2008), dendritic spine density (Beeson & Meitzen, 2023) and synaptic plasticity (Yankova et al., 2001). In female Syrian hamsters, E2 administration decreased dendritic spine density and altered cell morphology in the nucleus accumbens (as part of the ventral striatum) (Staffend et al., 2011), supporting the idea of localised E2-driven structural remodelling in the striatum.

It is well known that E2 can exert rapid effects via both rapid, non-genomic pathways through membrane-bound G-protein-coupled oestrogen receptors (within minutes) and genomic pathways, such as transcriptional regulation via nuclear oestrogen receptors (ERs) (hours to days) (Srivastava et al., 2013). ER subtypes are widely expressed in the brain, including in the striatum (Almey et al., 2015; Krentzel et al., 2021), in place for E2 to locally modulate neural plasticity. Our findings add evidence that such changes can occur rapidly after 24h and 12mg of oestradiol valerate administration in humans. The volumetric changes may reflect E2-induced alterations in neuronal morphology, dendritic spine density and shifts in cellular composition (neuron to glia cell ratio) (Giacobbo et al., 2022; Han et al., 1989; Rosen & Williams, 2001). E2 appears to affect different spine types in distinct ways: while spines of pyramidal neurons in the CA1 region of rodent hippocampus increase with E2 administration, certain spine subtypes in the nucleus accumbens were found to be reduced (Staffend et al., 2011). This synaptic pruning may contribute to reduced striatal volumes (Han et al., 1989, Giacobbo et al., 2022; Rosen & Williams, 2011; Mimura et al., 2003). GMV largely reflects cell bodies (somas) (Mercadante & Tadi, 2025). Reductions in striatal volumes have previously been observed with an increase in neuron to glia cell ratio in rats (Han et al., 1989). Given that neurons and glia cells differ in soma size (Purves et al., 2001) and that both neurons and glia cells express ERs (Donahue et al., 2000), such shifts in cellular composition may partly explain the striatal GMV decrease observed in our study.

Despite finding significant effects of E2 on striatum GMV we did not observe an effect for ACC, amygdala or hippocampus. This somehow contrasts previous findings showing that ACC volume is negatively correlated with E2 levels during the mid-luteal menstrual cycle phase (De Bondt et al., 2013). Other studies have reported plasticity of amygdala and hippocampus across different menstrual cycle phases (Lisofsky et al., 2015; Ossewaarde et al., 2010; Pletzer et al., 2018; Protopopescu et al., 2008; Zsido et al., 2023). This discrepancy is most likely explained by our focus on the effects of rapid E2 increase and speculatively, a different ratio of ER subtypes. Adult striatal neurons seem to primarily express G-protein-coupled ERs (Almey et al., 2022), whereas cells of the amygdala mainly express nuclear ERs (Österlund & Hurd, 2001). This receptor profile may allow for faster, non-genomic signalling cascades, and can account for the fast and immediate effects of E2 we observed in the striatum, compared to slower, genomic-driven changes. Thus, the striatum seems to have a heightened sensitivity to E2 and rapidly adapts in response to hormonal shifts. Other brain regions might adapt to E2 variations in a slower fashion due to their receptor profile. The striatum is a key subcortical structure involved in stress reactivity, emotion regulation, and reward processing (Etkin et al., 2015; Kogler et al., 2015). The dorsal striatum comprising the dorsal caudate and dorsal putamen, receives afferents from sensorimotor cortex and association cortices (Di Martino et al., 2008), supporting cognitive control of emotion, facilitating goal-directed modulation of affective responses (Etkin et al., 2015). The ventral striatum, including the nucleus accumbens and ventral parts of the caudate and putamen, is primarily connected to limbic and orbitofrontal regions (Di Martino et al., 2008), enabling evaluation of emotional valence and reward-related processing (Etkin et al., 2015) and is modulated by E2 through enhanced dopaminergic signalling (Bayer et al., 2020). The caudoventral striatum plays a critical role in integrating affective and motivational signals based on its functional and anatomical connections (Fudge & Haber, 2002). Thus, taken together with our data this suggest that the striatum is a critical region sensitive to E2-fluctations, and it is further driving motoric, senso-motoric, affective, stress and reward-related behaviour.

### 4.2. Association between emotion regulation and brain volume

Our findings indicate that, the tendency to use reappraisal or rumination did not impact GMV differently in phases of elevated vs. low E2 levels. Nevertheless, trait use of reappraisal was negatively associated with striatal GMV, independent of whether E2 levels remained low or increased rapidly. No association occurred between GMV and trait use of rumination.

Consistent with our results, Yoon and Jung (2025) reported that greater use of reappraisal was related to smaller GMV in the left ventral striatum in a mixed sex sample (not controlling for cycle phase or hormonal contraceptive use). Together these findings support that trait use of reappraisal can contribute to experience-dependent brain plasticity through use of specific cognitive and behavioural patterns (Giuliani et al., 2011). More broadly, individual differences in emotion regulation traits have been associated with structural variations in relevant brain areas (Giuliani et al., 2011 Kühn et al., 2012).

Taken together with our data, this suggests that also affective traits can shape brain anatomy. In our sample, participants reported more frequent use of reappraisal and less rumination compared to a predominantly female sample reported by Idzadpanah et al. (2017), potentially explaining the absence of an association between rumination and GMV. Importantly, our sample consisted of healthy young females, with participants excluded in case of moderate to severe depressive symptoms (BDI-II > 20). Prior studies linking rumination to GMV differences often involved clinical populations with higher and more dysfunctional rumination levels (Hilbert et al., 2015; Nolen-Hoeksema, 2000). Thus, the low frequency in rumination in the current sample might further contribute to the lack of effects for rumination.

Importantly, the impact of trait use of reappraisal on striatal GMV is independent of E2 levels. Thus, E2 levels do not shape striatal volumes differently in individuals using more reappraisal compared to those who apply less reappraisal in everyday life.

### 4.3. Limitations and future directions

By focusing on healthy, naturally cycling females in the early follicular phase of the menstrual cycle, our study was designed to isolate rapid E2-related structural effects while minimizing potential confounding variables (including progesterone levels). This targeted approach gains insight in the effects of E2 in healthy females, however, it does not capture the broader hormonal context in which sex hormones may naturally (inter-)act (Graham et al., 2018; Zsido et al., 2023). By applying a within subject design and controlling for baseline E2 levels (E2Δ), we tried to mitigate some of the naturally occurring hormone fluctuations. Hormonal levels may vary from cycle to cycle depending on the number of antral follicles recruited at the very beginning of the cycle. We also need to consider the individual differences in sensitivity to E2 as metabolic rates may introduce variability in E2 levels across participants. Oral administration of E2-valerate, as administered in the current study, undergoes first-pass metabolism in the gut and liver before it becomes bio-available (Gravelsins & Galea, 2025). Thus, further studies should explore the effect of different routes of E2 administration on GMV.

#### Potential clinical implications

As E2 administration can lead to rapid reductions in GMV volume, this may pose potential implications for mood and mental health. Given that smaller striatal volumes have been linked to better mental health outcomes (Dennison et al., 2015; Hilbert et al., 2015), such changes may represent a beneficial neuroplastic response—particularly during hormonally sensitive periods. There is some evidence that E2 treatment reduces perimenopausal depressive symptoms (transdermal administration) (Soares et al., 2001), improves mood in females with a history of major depressive disorder (Albert et al., 2021), and reduces anxiety- and stress-related behaviours in both animal models (Walf & and Frye, 2007) and naturally cycling females (Nouri et al., 2022). This emphasizing the importance of extending research to diverse female populations, including those diagnosed with affective disorders such as premenstrual dysphoric disorder (PMDD), where impaired emotion regulation and lower premenstrual E2 levels are associated with greater anxiety and stress (Yen et al., 2018). Since onset and relapse in females are often related to hormonal fluctuations across the menstrual cycle, pregnancy, the postpartum period or the transition to menopause (known as perimenopause) (Soares & Zitek, 2008) understanding individual E2-related brain changes could enable more targeted interventions and turning a window of vulnerability into a one of opportunity. Thus, further research should examine whether females with greater neuroplasticity are also more sensitive to hormonal fluctuations, and whether E2-induced striatal volume changes can predict symptom occurrence in those with affective disorders. Such insights could help to refine personalised hand hormone informed treatments to improve quality of life of females across the life span.

### 4.4. Conclusion

In summary, this study highlights the rapid effects of oestradiol (E2) and the influence of trait reappraisal use on striatal matter volume (GMV) in healthy, naturally cycling females. We demonstrated that decreased striatal volume is associated both with elevated E2 levels and with the use of reappraisal. This is the first study to assess rapid anatomical changes in the striatum and limbic system resulting from an E2 increase using a within-subject longitudinal design. The observed reductions in striatal volume after a rapid E2 increase in our study prove fast and efficient neuroplastic adaptations due to varying sex hormone levels in healthy females. As structural changes may serve as proxies for functional (Qing & Gong, 2016) and behavioural shifts (Wang et al., 2022), morphological changes of the striatum can subsequently signify meaningful alterations in circuitries associated with emotion regulation and mental health. In this context, E2 may foster structural refinement in key affective circuits, and may support mental health across female hormonal transition phases.

## Data Availability

All data produced in the present study are available upon reasonable request to the authors

## Author contributions

**AFD:** Formal analysis; Data visualisation; Data curation; Roles/Writing - original draft & review. **ER:** Data acquisition; Roles/Writing - review & editing. **ISP:** Conceptualization; Funding acquisition; Project administration; Supervision; Roles/Writing - review & editing. **BD:** Conceptualization; Funding acquisition; Project administration; Resources; Supervision; Roles/Writing - review & editing. **LK:** Conceptualization; Investigation; Data curation; Methodology; Project administration; Formal analysis; Supervision; Validation; Visualization; Roles/Writing – original draft, review & editing

## Declaration of AI use

During the preparation of this work the author(s) used ChatGPT (OpenAI, San Francisco, CA) to assist with language editing. After using this tool/service, the author(s) reviewed and edited the content as needed and take(s) full responsibility for the content of the publication.

