## Supplement for "Oestradiol, emotion regulation and the limbic system: effects on grey matter volume"

Supplementary methods

The regions of interest (ROI) - anterior cingulate cortex (ACC), hippocampus, and striatum – where further dissected into subregions including dorsal, subgenual and pregenual ACC, para- and hippocampus, ventral, and caudoventral striatum (Fig. S1)

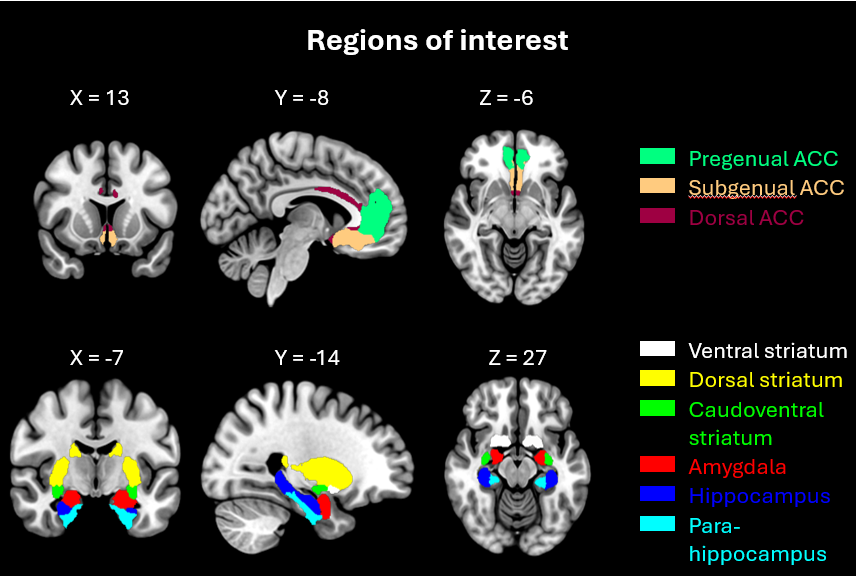

**Fig. S1 | Extended region of interest (ROI) in MNI-space**.

Supplementary figures & tables

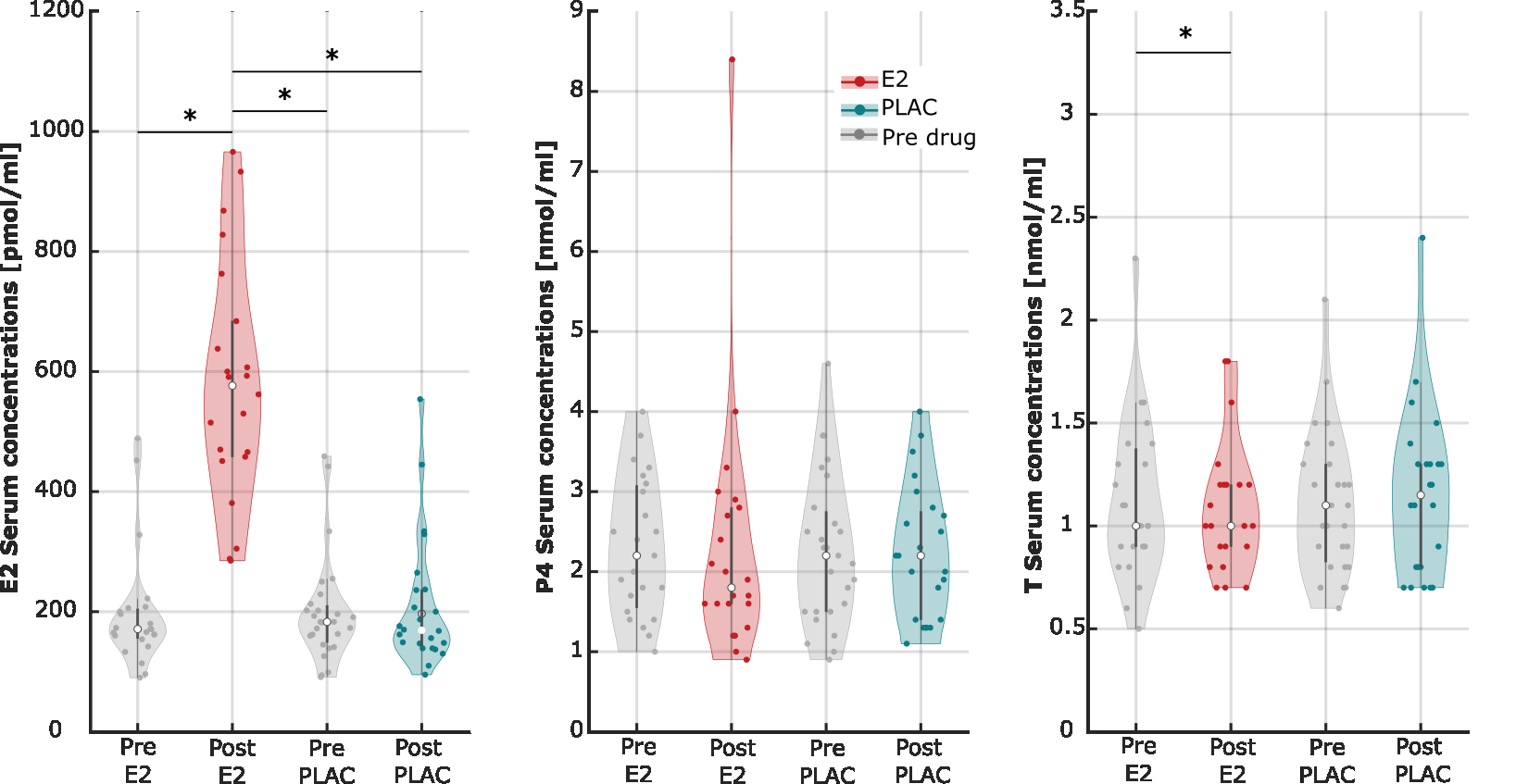

**Fig. S2 | Serum hormone concentrations of E2 [pmol/l], progesterone (P4) [nmol/l], testosterone (T) [nmol/l]** pre (grey) and post E2 (red) and post PLAC (blue) administration. Significance is indicated with an asterisk.

**Tab. S1 | Median serum hormone concentration of** E2 [pmol/l], P4 [nmol/l], T [nmol/l] pre and post drug administration (E2/PLAC) and significance levels of E2, P4 and T concentrations comparing drug conditions (Z-statistics, effect size). Significant differences are labelled with an asterisk.

|  | **PRE E2**  Median *(IQR)*  p *(Z,r)* | | | | **POST E2** Median *(IQR)*  p *(Z,r)* | **PRE PLAC**  Median *(IQR)*  p *(Z,r)* | **POST PLAC** Median *(IQR)*  p *(Z,r)* |
| --- | --- | --- | --- | --- | --- | --- | --- |
| **E2 [pmol/l]** | **171.0**  ***(155.5, 204.5)*** | | | | **576.5**  ***(458.0, 684.0)*** | **183.0**  ***(148.75, 210.25)*** | **169.0**  ***(143.9, 236.5)*** |
| PRE E2: | | | |  | < .001*  *(-4.02, -.88)* | .867  *(.17, .03)* | .689  *(-.40, -.09)* |
| POST E2: | | | |  |  | < .001*  *(4.12, .84)* | < .001*  *(3.82, .88)* |
| PRE PLAC: | | | |  |  |  | .145  *(1.46, .30)* |
| **P4 [nmol/l]** | **2.2**  ***(1.55, 3.075)*** | | | | **1.8**  ***(1.6, 2.8)*** | **2.2**  ***(1.5, 2.75)*** | **2.2**  ***(1.4, 2.75)*** |
| PRE E2: | | |  | | .199  *(1.28, .28)* | .961  *(-.05, -.01)* | .903  *(-.12, -.03)* |
| POST E2: | | |  | |  | .197  *(-1.29, -.27)* | .409  *(-.83, -.19)* |
| PRE PLAC: | | |  | |  |  | .379  (.88, .18) |
| **T [nmol/l]** | **1.0**  ***(0.9, 1.375)*** | | | | **1.0**  ***(0.9, 1.2)*** | **1.1**  ***(0.825, 1.3)*** | **1.15**  ***(0.8, 1.3)*** |
| PRE E2: | |  | | | .038*  *(2.07, .45)* | .734  *(-.34, -07)* | .536  *(-.62, -.14)* |
| POST E2: | |  | | |  | .134  *(-1.50, -.32)* | .134  *(-1.50, -.34)* |
| PRE PLAC: | |  | | |  |  | .238  *(-1.18, -.24)* |

**Tab. S2 | Median E2 increase in** [pmol/l] and centred log transformed values for E2 and PLAC drug condition. Inter quartile range is indicated.

| **E2 adminsitration** | | **PLAC administration** | |
| --- | --- | --- | --- |
| ΔE2 [pmol/l] | Centred & log  ΔE2 [log(pmol/l)] | ΔE2 [pmol/l] | Centred & log  ΔE2 [log(pmol/l)] |
| 576.5 *[458, 684]* | .050 *[-.180, .221]* | 1.69 *[143, 236.5]* | -.115 *[-.282, .222]* |

Effects of E2 increase on limbic GMV

**Tab. S3 | Statistical parameters from robust mixed linear regression analysing the relationship between oestradiol (E2) increase and regional gray matter volume (GMV) under E2 and placebo (PLAC) conditions**. Slope coefficients (β), standard errors (SE), t-statistics, and false discovery rate (FDR)-corrected p-values for the association between E2 increase and GMV across both drug conditions are reported. Additionally, p-values from Z-transformed slope comparisons (E2 vs. PLAC) are included to assess condition-specific differences in association strength. Significant associations (p < .05) are marked with an asterisk (*). Under the E2 condition, E2 increase was significantly negatively associated with GMV in the right ventral and caudoventral striatum, as well as bilaterally in the dorsal striatum, as well as overall striatum. These associations were not observed during the PLAC condition, and slope comparisons between conditions indicated significant differences in these regions.

| **Left amygdala** | | | | | | | | | |
| --- | --- | --- | --- | --- | --- | --- | --- | --- | --- |
|  | **E2 condition** | | | | **PLAC condition** | | | | **Z test** |
|  | β | SE | t-stat | p*_FDR_* | β | SE | t-stat | p*_FDR_* | p |
| **ΔE2** | -.028 | .067 | -.423 | .887 | .017 | .024 | .719 | .721 | .523 |
| **TIV** | .001 | <.001 | 5.172 | <.001* | .001 | < .001 | 4.917 | <.001* | .873 |
| **Age** | -.003 | .009 | -.339 | .887 | -.003 | .011 | -.256 | .721 | .977 |
| **Model** | N = 21, R² = .119, F = 10.5,  p < .001* | | | | N = 24, R² = .562, F= 8.6,  p < .001* | | | |  |
| **Right amygdala** | | | | | | | | | |
|  | **E2 condition** | | | | **PLAC condition** | | | | **Z test** |
|  | β | SE | t-stat | p*_FDR_* | β | SE | t-stat | p*_FDR_* | p |
| **ΔE2** | -.081 | .079 | -1.019 | .645 | .020 | .025 | .0804 | .721 | .226 |
| **TIV** | .002 | <.001 | 5.165 | >.001* | .002 | <.001 | 5.662 | <.001* | .902 |
| **Age** | -.001 | .011 | -.114 | .911 | -.006 | .012 | -.511 | .738 | .777 |
| **Model** | N = 21, R² = .658, F = 10.9,  p < .001* | | | | N = 24, R² = .634, F = 11.5,  p < .001* | | | |  |
| **Left ACC** | | | | | | | | | |
|  | **E2 condition** | | | | **PLAC condition** | | | | **Z test** |
|  | β | SE | t-stat | p*_FDR_* | β | SE | t-stat | p*_FDR_* | p |
| **ΔE2** | -.130 | .266 | -.487 | .632 | .005 | .083 | .054 | .957 | .630 |
| **TIV** | .007 | .001 | 6.206 | <.001* | .006 | .001 | 6.681 | <.001* | .843 |
| **Age** | -.077 | -.039 | -2.011 | .121 | -.073 | .039 | -1.859 | .156 | .933 |
| **Model** | N = 21, R² = .763, F = 18.3,  p < .001* | | | | N = 24, R² = .733, F = 18.3,  p < .001* | | | |  |
| **Right ACC** | | | | | | | | | |
|  | **E2 condition** | | | | **PLAC condition** | | | | **Z test** |
|  | β | SE | t-stat |  | β | SE | t-stat |  | p |
| **ΔE2** | -.156 | .268 | -.058 | .632 | -.038 | .070 | -.558 | .699 | .675 |
| **TIV** | .006 | .001 | 5.930 | <.001* | .006 | .001 | 7.836 | <.001* | .961 |
| **Age** | -.035 | .039 | -.906 | .567 | -.028 | .033 | -.839 | .617 | .888 |
| **Model** | N = 21, R² = .721, F = 14.7,  p < .001* | | | | N = 24, R² = .772, F = 22.6,  p < .001* | | | |  |
| **Left perigenual ACC** | | | | | | | | | |
|  | **E2 condition** | | | | **PLAC condition** | | | | **Z test** |
|  | β | SE | t-stat | p*_FDR_* | β | SE | t-stat | p*_FDR_* | p |
| **ΔE2** | .015 | .190 | .078 | .939 | -.001 | .063 | -.017 | .987 | .937 |
| **TIV** | .005 | < .001 | 6.041 | <.001* | .004 | < .001 | 5.972 | <.001* | .777 |
| **Age** | -.059 | .027 | -2.132 | .096 | -.057 | .030 | -1.904 | .143 | .962 |
| **Model** | N = 21, R² = .752, F = 17.2,  p < .001* | | | | N = 24, R² = .694, F = 15.1,  p < .001* | | | |  |
| **Right perigenual ACC** | | | | | | | | | |
|  | **E2 condition** | | | | **PLAC condition** | | | | **Z test** |
|  | β | SE | t-stat | p*_FDR_* | β | SE | t-stat | p*_FDR_* | p |
| **ΔE2** | -.045 | .194 | -.235 | .939 | -.033 | .0517 | -.642 | .634 | .208 |
| **TIV** | .004 | .001 | 5.450 | <.001* | 0.004 | 0.001 | 7.1257 | <.001* | .709 |
| **Age** | -.028 | .028 | -.996 | .500 | -.023 | .024 | -.932 | .543 | 1 |
| **Model** | N = 21, R² = .686, F = 12.4,  p < .001* | | | | N = 24, R² = .741, F = 19.1,  p < .001* | | | |  |
| **Left subgenual ACC** | | | | | | | | | |
|  | **E2 condition** | | | | **PLAC condition** | | | | **Z test** |
|  | β | SE | t-stat | p*_FDR_* | β | SE | t-stat | p*_FDR_* | p |
| **ΔE2** | -.111 | .087 | -1.27 | .330 | .006 | .025 | .246 | .936 | .196 |
| **TIV** | .002 | < .001 | 4.686 | .001* | .002 | <.001 | 5.305 | <.001* | .823 |
| **Age** | -.013 | .013 | -1.025 | .379 | -.012 | .012 | -1.018 | .530 | .966 |
| **Model** | N = 21, R² = .623, F = 10.6,  p < .001* | | | | N = 24, R² = .617, F = 10.7,  p < .001* | | | |  |
| **Right subgenual ACC** | | | | | | | | | |
|  | **E2 condition** | | | | **PLAC condition** | | | | **Z test** |
|  | β | SE | t-stat | p*_FDR_* | β | SE | t-stat | p*_FDR_* | p |
| **ΔE2** | -.087 | .065 | -1.336 | .330 | -.002 | .019 | -.081 | .936 | .623 |
| **TIV** | .002 | <.001 | 6.123 | <.001* | .002 | <.001 | 6.792 | <.001* | .743 |
| **Age** | -.008 | 1 | -.904 | .379 | -.009 | .009 | -.951 | .530 | .674 |
| **Model** | N = 21, R² = .747, F = 16.8,  p < .001* | | | | N = 24, R² = .720, F = 17.1,  p < .001* | | | |  |
| **Left dorsal ACC** | | | | | | | | | |
|  | **E2 condition** | | | | **PLAC condition** | | | | **Z test** |
|  | β | SE | t-stat | p*_FDR_* | β | SE | t-stat | p*_FDR_* | p |
| **ΔE2** | -.028 | .027 | -1.040 | .469 | .002 | .008 | .296 | .771 | .280 |
| **TIV** | < .001 | <.001 | 4.119 | .002* | <.001 | <.001 | 5.241 | <.001* | .845 |
| **Age** | -.007 | .004 | -1.74 | .198 | -.006 | .004 | -1.533 | .282 | .839 |
| **Model** | N = 21, R² = .628, F = 9.6,  p < .001* | | | | N = 24, R² = .628, F = 11.3,  p < .001* | | | |  |
| **Right dorsal ACC** | | | | | | | | | |
|  | **E2 condition** | | | | **PLAC condition** | | | | **Z test** |
|  | β | SE | t-stat | p*_FDR_* | β | SE | t-stat | p*_FDR_* | p |
| **ΔE2** | .015 | .037 | .393 | .792 | -.005 | .012 | -.382 | .771 | .984 |
| **TIV** | <.001 | <.001 | 4.093 | .003* | <.001 | <.001 | 3.947 | .002* | .595 |
| **Age** | -.001 | .005 | -.267 | .792 | .002 | .006 | .326 | .771 | .948 |
| **Model** | N = 21, R² = .522, F = 6.2,  p = .005* | | | | N = 24, R² =.445, F = 5.3,  p =.007* | | | |  |
| **Left total hippocampus** | | | | | | | | | |
|  | **E2 condition** | | | | **PLAC condition** | | | | **Z test** |
|  | β | SE | t-stat | p*_FDR_* | β | SE | t-stat | p*_FDR_* | p |
| **ΔE2** | .029 | .191 | .153 | .880 | -.084 | .051 | 1.666 | .223 | .780 |
| **TIV** | .003 | .001 | 4.011 | .003* | .003 | .001 | 4.946 | .001* | .823 |
| **Age** | -.008 | .028 | -.293 | .880 | -.013 | .024 | -.533 | .600 | .899 |
| **Model** | N = 21, R² = .516, F = 6.0,  p = .005* | | | | N = 24, R² = .587, F= 9.5,  p < .001* | | | |  |
| **Right total hippocampus** | | | | | | | | | |
|  | **E2 condition** | | | | **PLAC condition** | | | | **Z test** |
|  | β | SE | t-stat | p*_FDR_* | β | SE | t-stat | p*_FDR_* | p |
| **ΔE2** | .044 | .197 | .223 | .880 | .040 | .054 | .731 | .600 | .983 |
| **TIV** | .003 | .001 | 3.958 | .003* | .003 | .001 | 4.569 | .001* | .765 |
| **Age** | -.015 | .029 | -.528 | .880 | -.014 | .026 | -.553 | .600 | .981 |
| **Model** | N = 21, R² = .518, F = 6.1,  p = .005* | | | | N = 24, R² = .535, F= 7.7,  p = .001* | | | |  |
| **Left hippocampus** | | | | | | | | | |
|  | **E2 condition** | | | | **PLAC condition** | | | | **Z test** |
|  | β | SE | t-stat | p*_FDR_* | β | SE | t-stat | p*_FDR_* | p |
| **ΔE2** | .014 | .095 | .148 | .845 | .037 | .027 | 1.385 | .330 | .813 |
| **TIV** | .002 | <.001 | 4.963 | .056* | .002 | <.001 | 5.076 | .002* | .511 |
| **Age** | -.004 | .014 | -.317 | .845 | -.012 | .013 | -1.041 | .771 | .634 |
| **Model** | N = 21, R² = .620, F = 9.2,  p < .001* | | | | N = 24, R² = .604, F = 10.2,  p < .001* | | | |  |
| **Right** **hippocampus** | | | | | | | | | |
|  | **E2 condition** | | | | **PLAC condition** | | | | **Z test** |
|  | β | SE | t-stat | p*_FDR_* | β | SE | t-stat | p*_FDR_* | p |
| **ΔE2** | .016 | .119 | .134 | .972 | .013 | .035 | .379 | .851 | .921 |
| **TIV** | .002 | <.001 | 3.942 | .003* | .002 | <.001 | 3.829 | .003* | .942 |
| **Age** | <.001 | .0172 | .035 | .972 | <.001 | .017 | -.058 | .954 | .883 |
| **Model** | N = 21, R² = .497, F = 5.6,  p = .007* | | | | N = 24, R² = .433, F = 5.1,  p = .009* | | | |  |
| **Left para-hippocampus** | | | | | | | | | |
|  | **E2 condition** | | | | **PLAC condition** | | | | **Z test** |
|  | β | SE | t-stat | p*_FDR_* | β | SE | t-stat | p*_FDR_* | p |
| **ΔE2** | .025 | .112 | .224 | .671 | .042 | .029 | 1.443 | .353 | .884 |
| **TIV** | .001 | <.001 | 2.693 | <.001* | .001 | <.001 | 4.049 | <.001* | .809 |
| **Age** | -.003 | .016 | -.198 | .688 | .004 | .014 | .295 | .678 | .733 |
| **Model** | N = 21, R² = .323, F = 2.7,  p = .078 | | | | N = 24, R² = .482, F = 6.2,  P = .004* | | | |  |
| **Right para-hippocampus** | | | | | | | | | |
|  | **E2 condition** | | | | **PLAC condition** | | | | **Z test** |
|  | β | SE | t-stat | p*_FDR_* | β | SE | t-stat | p*_FDR_* | p |
| **ΔE2** | .036 | .094 | .384 | .615 | .026 | .027 | .98 | .678 | .387 |
| **TIV** | .001 | <.001 | 3.391 | <.001* | .001 | <.001 | 4.307 | <.001* | .675 |
| **Age** | -.014 | .014 | -1.027 | .627 | -.011 | .013 | -.893 | .634 | .968 |
| **Model** | N = 21, R² = .472, F = 5.1,  p = .011* | | | | N = 24, R² = .521, F = 7.2,  p = .002* | | | |  |
| **Left striatum** | | | | | | | | | |
|  | **E2 condition** | | | | **PLAC condition** | | | | **Z test** |
|  | β | SE | t-stat | p*_FDR_* | β | SE | t-stat | p*_FDR_* | p |
| **ΔE2** | -.685 | .247 | -2.769 | .026* | .009 | .078 | .117 | .908 | .008* |
| **TIV** | .005 | .001 | 5.410 | .001* | .007 | .001 | 7.413 | < .001* | .353 |
| **Age** | -.027 | .036 | .749 | .696 | -.011 | .037 | -.301 | .908 | .759 |
| **Model** | N = 21, R² = .742, F = 16.3,  p < .001* | | | | N = 24, R² = .745, F= 19.5,  p < .001* | | | |  |
| **Right** **striatum** | | | | | | | | | |
|  | **E2 condition** | | | | **PLAC condition** | | | | **Z test** |
|  | β | SE | t-stat | p*_FDR_* | β | SE | t-stat | p*_FDR_* | p |
| **ΔE2** | -.854 | .236 | -3.625 | .004* | -.009 | .085 | -.100 | .921 | <.001* |
| **TIV** | .006 | .001 | 6.334 | < .001* | .007 | .001 | 6.857 | < .001* | .604 |
| **Age** | -.021 | .034 | -6.09 | .661 | -.043 | .040 | -1.079 | .446 | .667 |
| **Model** | N = 21, R² = .803, F = 23.1,  p < .001* | | | | N = 24, R² = .729, F= 17.9,  P < .001* | | | |  |
| **Left ventral striatum** | | | | | | | | | |
|  | **E2 condition** | | | | **PLAC condition** | | | | **Z test** |
|  | β | SE | t-stat | p*_FDR_* | β | SE | t-stat | p*_FDR_* | p |
| **ΔE2** | -.100 | .045 | -2.228 | .060 | -.009 | .015 | -.584 | .576 | .052 |
| **TIV** | <.001 | <.001 | 4.103 | .002* | <.001 | <.001 | 4.896 | .001* | .766 |
| **Age** | -.004 | .007 | -.632 | .643 | -.004 | .007 | -.569 | .576 | .982 |
| **Model** | N = 21, R² = .631, F = 9.7,  p < .001* | | | | N = 24, R² = .575, F = 9.0,  p< .001* | | | |  |
| **Right** **ventral striatum** | | | | | | | | | |
|  | **E2 condition** | | | | **PLAC condition** | | | | **Z test** |
|  | β | SE | t-stat | p*_FDR_* | β | SE | t-stat | p*_FDR_* | p |
| **ΔE2** | -.110 | .036 | -3.099 | .013* | -.016 | .016 | -1.044 | .464 | .016* |
| **TIV** | <.001 | <.001 | 5.721 | <.001 | <.001 | <.001 | 4.423 | <.001 | .897 |
| **Age** | -.002 | .005 | -.358 | .725 | -0.009 | 0.007 | -1.218 | .464 | .429 |
| **Model** | N = 21, R² = .760, F = 18.0,  p < .001* | | | | N = 24, R² = .569, F = 8.8,  p < .001* | | | |  |
| **Left dorsal striatum** | | | | | | | | | |
|  | **E2 condition** | | | | **PLAC condition** | | | | **Z test** |
|  | β | SE | t-stat | p*_FDR_* | β | SE | t-stat | p*_FDR_* | p |
| **ΔE2** | -.584 | .226 | -2.592 | .029* | .009 | .067 | .138 | .892 | .012* |
| **TIV** | .004 | <.001 | 4.669 | >.001* | .005 | <.001 | 7.082 | <.001* | .300 |
| **Age** | -.033 | .033 | -.995 | .393 | -.011 | .032 | -.359 | .869 | .644 |
| **Model** | N = 21, R² = .699, F = 13.1,  p < .001* | | | | N = 24, R² = .729, F = 18.0,  p < .001* | | | |  |
| **Right dorsal striatum** | | | | | | | | | |
|  | **E2 condition** | | | | **PLAC condition** | | | | **Z test** |
|  | β | SE | t-stat | p*_FDR_* | β | SE | t-stat | p*_FDR_* | p |
| **ΔE2** | -.073 | .211 | -3.465 | .006* | -.029 | .075 | -.389 | .869 | .002* |
| **TIV** | .005 | <.001 | 5.696 | <.001* | .005 | <.001 | 6.312 | <.001* | .615 |
| **Age** | -.027 | .031 | -.877 | .393 | -.070 | 0.036 | -1.965 | .127 | .359 |
| **Model** | N = 21, R² = .778, F = 19.8,  p < .001* | | | | N = 24, R² = .430, F = 17.1,  p < .001* | | | |  |
| **Left caudoventral striatum** | | | | | | | | | |
|  | **E2 condition** | | | | **PLAC condition** | | | | **Z test** |
|  | β | SE | t-stat | p*_FDR_* | β | SE | t-stat | p*_FDR_* | p |
| **ΔE2** | -.022 | .016 | -1.338 | .298 | .006 | .004 | 1.208 | .242 | .097 |
| **TIV** | <.001 | y.001 | 3.996 | .006* | <.001 | <.001 | 5.209 | >.001* | .995 |
| **Age** | <.001 | .002 | -.068 | .947 | -.002 | .002 | -1.009 | .325 | .539 |
| **Model** | N = 21, R² = .560, F = 7.2,  p < .001* | | | | N = 24, R² = .615, F = 10.6,  p < .001* | | | |  |
| **Right caudoventral striatum** | | | | | | | | | |
|  | **E2 condition** | | | | **PLAC condition** | | | | **Z test** |
|  | β | SE | t-stat | p*_FDR_* | β | SE | t-stat | p*_FDR_* | p |
| **ΔE2** | -.044 | 0.015 | -2.831 | .023* | .009 | .005 | 1.701 | .209 | .001* |
| **TIV** | <.001 | <.001 | 2.972 | .023* | <.001 | <.011 | 5.059 | <.001* | .199 |
| **Age** | <.001 | .002 | -.429 | .808 | -.003 | .002 | -1.320 | .242 | .502 |
| **Model** | N = 21, R² = .567, F = 7.4,  p =.002* | | | | N = 24, R² = .616, F = 10.7,  p < .001* | | | |  |

Effects of trait emotion regulation on limbic GMV

### Reappraisal

**Tab. S4 | Statistical parameters from robust mixed linear regression analysing the relationship between emotion regulation strategy (trait reappraisal and rumination) and regional gray matter volume (GMV) under E2 and placebo (PLAC) conditions**. Slope coefficients (β), standard errors (SE), t-statistics, and false discovery rate (FDR)-corrected p-values for the association between E2 increase and GMV across both drug conditions are reported. Additionally, p-values from Z-transformed slope comparisons (E2 vs. PLAC) are included to assess condition-specific differences in association strength. Significant associations (p < .05) are marked with an asterisk (*). Trait reappraisal was negatively associated with right ventral and bilateral caudoventral and dorsal striatum GMV. However, slopes did not differ between E2 and PLAC drug condition.

| **Left amygdala** | | | | | | | | | | | | | | | | | | | | | | | | | | | | | | | | | |
| --- | --- | --- | --- | --- | --- | --- | --- | --- | --- | --- | --- | --- | --- | --- | --- | --- | --- | --- | --- | --- | --- | --- | --- | --- | --- | --- | --- | --- | --- | --- | --- | --- | --- |
|  | | | **E2 condition** | | | | | | | | | | | | | | | **PLAC condition** | | | | | | | | | | | | | | | **Z test** |
|  | | | | | | β | | | SE | | | t-stat | | | p*_FDR_* | | | β | | | | | | SE | | | t-stat | | | p*_FDR_* | | | p |
| **Reappraisal** | | | | | | -.003 | | | .009 | | | -.370 | | | .715 | | | -.014 | | | | | | .010 | | | -1.422 | | | .337 | | | .443 |
| **TIV** | | | | | | .001 | | | <.001 | | | 5.386 | | | <.001* | | | .001 | | | | | | <.001 | | | 5.086 | | | <.001* | | | .939 |
| **Age** | | | | | | .005 | | | .008 | | | .655 | | | .715 | | | .006 | | | | | | .009 | | | .751 | | | .559 | | | .932 |
| **Model** | | | | | | N = 27, R² = .561, F = 9.8,  p < .001* | | | | | | | | | | | | | | | N = 27, R² = .542, F= 9.1,  p < .001* | | | | | | | | | | | | |
| **Right amygdala** | | | | | | | | | | | | | | | | | | | | | | | | | | | | | | | | | |
|  | | | **E2 condition** | | | | | | | | | | | | | | | **PLAC condition** | | | | | | | | | | | | | | | **Z test** |
|  | | | | | | β | | | SE | | | t-stat | | | p*_FDR_* | | | β | | | | | | SE | | | t-stat | | | p*_FDR_* | | | p |
| **Reappraisal** | | | | | | -.004 | | | .011 | | | -.397 | | | .715 | | | -.006 | | | | | | .011 | | | -.593 | | | .559 | | | .886 |
| **TIV** | | | | | | .002 | | | <.001 | | | 5.555 | | | <.001* | | | .001 | | | | | | <.001 | | | 5.138 | | | <.001* | | | .806 |
| **Age** | | | | | | .017 | | | .010 | | | 1.199 | | | .486 | | | .007 | | | | | | .010 | | | .667 | | | .559 | | | .713 |
| **Model** | | | | | | N = 27, R² = .574, F = 10.3,  p < .001* | | | | | | | | | | | | | | | N = 27, R² = .538, F = 8.9,  p < .001* | | | | | | | | | | | | |
| **Left ACC** \| \| \| \| \| \| \| \| \| \| \|; \| **E2 condition** \| \| \| \| \| **PLAC condition** \| \| \| \| \| **Z test**; \| \| β \| SE \| t-stat \| p*_FDR_* \| β \| \| SE \| t-stat \| p*_FDR_* \| p; **Reappraisal** \| \| -.005 \| .039 \| -.140 \| .890 \| -.033 \| \| .039 \| -.860 \| .479 \| .938; **TIV** \| \| .006 \| .001 \| 5.545 \| <.001* \| .006 \| \| .001 \| 5.809 \| <.001* \| .915; **Age** \| \| -.051 \| .035 \| -1.449 \| .322 \| -.057 \| \| .035 \| -1.625 \| .236 \| .872; **Model** \| \| N = 27, R² = .626, F = 12.9, p < .001* \| \| \| \| \| N = 27, R² = .653, F= 14.5, p < .001* \| \| \| \| | | | | | | | | | | | | | | | | | | | | | | | | | | | | | | | | | |
| **Right ACC** \| \| \| \| \| \| \| \| \| \| \|; \| **E2 condition** \| \| \| \| \| **PLAC condition** \| \| \| \| \| **Z test**; \| \| β \| SE \| t-stat \| p*_FDR_* \| β \| \| SE \| t-stat \| p*_FDR_* \| p; **Reappraisal** \| \| -.023 \| .037 \| -.625 \| .924 \| -.036 \| \| .035 \| -1.045 \| .952 \| .889; **TIV** \| \| .006 \| .001 \| 5.873 \| .001* \| .006 \| \| .001 \| 6.370 \| .001* \| .844; **Age** \| \| -.016 \| .033 \| -.487 \| .924 \| -.020 \| \| .031 \| -.632 \| .952 \| .877; **Model** \| \| N = 27, R² = .624, F = 12.7, p < .001* \| \| \| \| \| N = 27, R² = .665, F = 15.2, p < .001* \| \| \| \| | | | | | | | | | | | | | | | | | | | | | | | | | | | | | | | | | |
| **Left perigenual ACC** | | | | | | | | | | | | | | | | | | | | | | | | | | | | | | | | | |
|  | | **E2 condition** | | | | | | | | | | | | | | | **PLAC condition** | | | | | | | | | | | | | | | **Z test** | |
|  | | | | | β | | | SE | | | t-stat | | | p*_FDR_* | | | β | | | | | | SE | | | t-stat | | | p*_FDR_* | | | p | |
| **Reappraisal** | | | | | -.001 | | | .037 | | | -.026 | | | .979 | | | -.019 | | | | | | .027 | | | -.698 | | | .492 | | | .635 | |
| **TIV** | | | | | .004 | | | .001 | | | 5.443 | | | <.001* | | | .004 | | | | | | .001 | | | 5.40 | | | <.001* | | | .893 | |
| **Age** | | | | | -.043 | | | .025 | | | -1.729 | | | .195 | | | -.045 | | | | | | .025 | | | -1.822 | | | .163 | | | .949 | |
| **Model** | | | | | N = 27, R² = .629, F = 13.0,  p < .001* | | | | | | | | | | | | | | | N = 27, R² = .648, F = 14.1,  p < .001* | | | | | | | | | | | | | |
| **Right perigenual ACC** | | | | | | | | | | | | | | | | | | | | | | | | | | | | | | | | | |
|  | | **E2 condition** | | | | | | | | | | | | | | | **PLAC condition** | | | | | | | | | | | | | | | **Z test** | |
|  | | | | | β | | | SE | | | t-stat | | | p*_FDR_* | | | β | | | | | | SE | | | t-stat | | | *p_FDR_* | | | p | |
| **Reappraisal** | | | | | -.017 | | | .026 | | | -.644 | | | .637 | | | -.025 | | | | | | .025 | | | -1.012 | | | .483 | | | .813 | |
| **TIV** | | | | | .004 | | | .001 | | | 5.532 | | | <.001* | | | .004 | | | | | | .001 | | | 5.958 | | | <.001* | | | .888 | |
| **Age** | | | | | -.015 | | | .024 | | | -.636 | | | .637 | | | -.016 | | | | | | .023 | | | -.718 | | | .492 | | | .969 | |
| **Model** | | | | | N = 27, R² = .601, F = 11.5,  p < .001* | | | | | | | | | | | | | | | N = 27, R² = .638, F = 13.5,  p < .001* | | | | | | | | | | | | | |
| **Left subgenual ACC** | | | | | | | | | | | | | | | | | | | | | | | | | | | | | | | | | |
|  | | **E2 condition** | | | | | | | | | | | | | | | **PLAC condition** | | | | | | | | | | | | | | | **Z test** | |
|  | | | | | β | | | SE | | | t-stat | | | p*_FDR_* | | | β | | | | | | SE | | | t-stat | | | p*_FDR_* | | | p | |
| **Reappraisal** | | | | | .002 | | | .013 | | | .168 | | | .868 | | | -.006 | | | | | | .012 | | | -.466 | | | .646 | | | .658 | |
| **TIV** | | | | | .001 | | | <.001 | | | 4.203 | | | .001* | | | .001 | | | | | | < .001 | | | 4.474 | | | .001* | | | .987 | |
| **Age** | | | | | -.005 | | | .011 | | | -.407 | | | .864 | | | -.009 | | | | | | .011 | | | -.809 | | | .641 | | | .796 | |
| **Model** | | | | | N = 27, R² = .462, F = 6.6,  p = .002* | | | | | | | | | | | | | | | N = 27, R² = .507, F = 7.9,  p < .001* | | | | | | | | | | | | | |
| **Right subgenual ACC** | | | | | | | | | | | | | | | | | | | | | | | | | | | | | | | | | |
|  | | **E2 condition** | | | | | | | | | | | | | | | **PLAC condition** | | | | | | | | | | | | | | | **Z test** | |
|  | | | | | β | | | SE | | | t-stat | | | p*_FDR_* | | | β | | | | | | SE | | | t-stat | | | p*_FDR_* | | | p | |
| **Reappraisal** | | | | | -.003 | | | .009 | | | -.363 | | | .864 | | | -.005 | | | | | | .009 | | | -.570 | | | .646 | | | .892 | |
| **TIV** | | | | | .001 | | | <.001 | | | 5.516 | | | <.001* | | | .001 | | | | | | <.001 | | | 5.764 | | | <.001* | | | .960 | |
| **Age** | | | | | -.004 | | | .008 | | | -.507 | | | .864 | | | -.008 | | | | | | .008 | | | -.992 | | | .641 | | | .744 | |
| **Model** | | | | | N = 27, R² = .595, F = 11.3,  p < .001* | | | | | | | | | | | | | | | N = 27, R² = .629, F = 13,  p < .001* | | | | | | | | | | | | | |
| **Left dorsal ACC** | | | | | | | | | | | | | | | | | | | | | | | | | | | | | | | | | |
|  | | **E2 condition** | | | | | | | | | | | | | | | **PLAC condition** | | | | | | | | | | | | | | | **Z test** | |
|  | | | | | β | | | SE | | | t-stat | | | p*_FDR_* | | | β | | | | | | SE | | | t-stat | | | p*_FDR_* | | | p | |
| **Reappraisal** | | | | | -.006 | | | .004 | | | -1.630 | | | .234 | | | -.008 | | | | | | .003 | | | -2.256 | | | .068 | | | .785 | |
| **TIV** | | | | | <.001 | | | <.001 | | | 4.209 | | | .002* | | | < .001 | | | | | | <.001 | | | 4.946 | | | <.001* | | | .902 | |
| **Age** | | | | | -.004 | | | .004 | | | -1.133 | | | .403 | | | -.004 | | | | | | .003 | | | -1.247 | | | .338 | | | .981 | |
| **Model** | | | | | N = 27, R² = .516, F = 8.2,  p < .001* | | | | | | | | | | | | | | | N = 27, R² = .601, F = 11.5,  p < .001* | | | | | | | | | | | | | |
| **Right dorsal ACC** | | | | | | | | | | | | | | | | | | | | | | | | | | | | | | | | | |
|  | | **E2 condition** | | | | | | | | | | | | | | | **PLAC condition** | | | | | | | | | | | | | | | **Z test** | |
|  | | | | | β | | | SE | | | t-stat | | | p*_FDR_* | | | β | | | | | | SE | | | t-stat | | | p*_FDR_* | | | p | |
| **Reappraisal** | | | | | -.002 | | | .005 | | | -.360 | | | .722 | | | -.003 | | | | | | .005 | | | -.602 | | | .553 | | | .864 | |
| **TIV** | | | | | <.001 | | | <.001 | | | 3.610 | | | .004* | | | .001 | | | | | | <.001 | | | 3.645 | | | .004* | | | .977 | |
| **Age** | | | | | .002 | | | .005 | | | .502 | | | .722 | | | .003 | | | | | | .005 | | | .607 | | | .553 | | | .940 | |
| **Model** | | | | | N = 27, R² = .365, F = 4.4,  p = .014* | | | | | | | | | | | | | | | N = 27, R² =.370, F = 4.5,  p =.013* | | | | | | | | | | | | | |
| **Left total hippocampus** \| \| \| \| \| \| \| \| \| \| \|; \| **E2 condition** \| \| \| \| \| **PLAC condition** \| \| \| \| \| **Z test**; \| \| β \| SE \| t-stat \| p*_FDR_* \| β \| \| SE \| t-stat \| p*_FDR_* \| p; **Reappriasal** \| \| .002 \| .026 \| .071 \| .944 \| -.002 \| \| .023 \| -.115 \| .952 \| .943; **TIV** \| \| .003 \| .001 \| 3.902 \| .002* \| .003 \| \| .001 \| 4.427 \| .001* \| .179; **Age** \| \| .018 \| .023 \| .762 \| .908 \| .014 \| \| .021 \| .670 \| .952 \| .858; **Model** \| \| N = 27, R² = .400, F = 5.1, p = .007* \| \| \| \| \| N = 27, R² = .462, F= 6.6, p = .002* \| \| \| \| | | | | | | | | | | | | | | | | | | | | | | | | | | | | | | | | | |
| **Right total hippocampus** \| \| \| \| \| \| \| \| \| \| \|; \| **E2 condition** \| \| \| \| \| **PLAC condition** \| \| \| \| \| **Z test**; \| \| β \| SE \| t-stat \| p*_FDR_* \| β \| \| SE \| t-stat \| p*_FDR_* \| p; **Reappraisal** \| \| .010 \| .024 \| .438 \| .757 \| -.001 \| \| .023 \| -.061 \| .460 \| .825; **TIV** \| \| .003 \| .001 \| 4.475 \| <.001* \| .003 \| \| .001 \| 4.368 \| <.001* \| .779; **Age** \| \| .006 \| .021 \| .296 \| .757 \| -.002 \| \| .020 \| -.123 \| .534 \| .752; **Model** \| \| N = 27, R² = .479, F = 7.0, p = .002* \| \| \| \| \| N = 27, R² = .472, F= 6.84, p = .002* \| \| \| \| | | | | | | | | | | | | | | | | | | | | | | | | | | | | | | | | | |
| **Left hippocampus** | | | | | | | | | | | | | | | | | | | | | | | | | | | | | | | | | |
|  | | **E2 condition** | | | | | | | | | | | | | | | **PLAC condition** | | | | | | | | | | | | | | | **Z test** | |
|  | | | | | β | | | SE | | | t-stat | | | p*_FDR_* | | | β | | | | | | SE | | | t-stat | | | p*_FDR_* | | | p | |
| **Reappraisal** | | | | | <.001 | | | .013 | | | -.066 | | | .948 | | | <-.001 | | | | | | .013 | | | -.023 | | | .982 | | | .976 | |
| **TIV** | | | | | .002 | | | <.001 | | | 4.690 | | | .001* | | | .002 | | | | | | >.001 | | | 4.364 | | | .001* | | | .774 | |
| **Age** | | | | | .009 | | | .012 | | | .771 | | | .819 | | | .006 | | | | | | .012 | | | .550 | | | .946 | | | .869 | |
| **Model** | | | | | N = 27, R² = .490, F = 7.4,  p = .001* | | | | | | | | | | | | | | | N = 27, R² = .457, F = 6.4,  p = .003* | | | | | | | | | | | | | |
| **Right hippocampus** | | | | | | | | | | | | | | | | | | | | | | | | | | | | | | | | | |
|  | | **E2 condition** | | | | | | | | | | | | | | | **PLAC condition** | | | | | | | | | | | | | | | **Z test** | |
|  | | | | | β | | | SE | | | t-stat | | | p*_FDR_* | | | β | | | | | | SE | | | t-stat | | | p*_FDR_* | | | p | |
| **Reappraisal** | | | | | .006 | | | .014 | | | .415 | | | .819 | | | .005 | | | | | | .014 | | | .359 | | | .946 | | | .965 | |
| **TIV** | | | | | .002 | | | <.001 | | | 4.470 | | | .001* | | | .001 | | | | | | <.001 | | | 4.000 | | | .002* | | | .706 | |
| **Age** | | | | | .007 | | | .012 | | | .556 | | | .819 | | | .003 | | | | | | .012 | | | .271 | | | .946 | | | .837 | |
| **Model** | | | | | N = 27, R² = .474, F = 6.9,  p = .002* | | | | | | | | | | | | | | | N = 27, R² = .424, F = 5.6,  p = .005* | | | | | | | | | | | | | |
| **Left para-hippocampus** | | | | | | | | | | | | | | | | | | | | | | | | | | | | | | | | | |
|  | | **E2 condition** | | | | | | | | | | | | | | | **PLAC condition** | | | | | | | | | | | | | | | **Z test** | |
|  | | | | | β | | | SE | | | t-stat | | | p*_FDR_* | | | β | | | | | | SE | | | t-stat | | | p*_FDR_* | | | p | |
| **Reappraisal** | | | | | .001 | | | .001 | | | .095 | | | .925 | | | -.002 | | | | | | .013 | | | -.013 | | | .898 | | | .875 | |
| **TIV** | | | | | .001 | | | <.001 | | | 2.994 | | | .020* | | | .001 | | | | | | < .001 | | | 3.707 | | | .004* | | | .789 | |
| **Age** | | | | | .011 | | | .013 | | | .861 | | | .797 | | | .009 | | | | | | .011 | | | .805 | | | .803 | | | .922 | |
| **Model** | | | | | N = 27, R² = .283, F = 3.0,  p = .050 | | | | | | | | | | | | | | | N = 27, R² = .374, F = 4.6,  p = .012* | | | | | | | | | | | | | |
| **Right para-hippocampus** | | | | | | | | | | | | | | | | | | | | | | | | | | | | | | | | | |
|  | | **E2 condition** | | | | | | | | | | | | | | | **PLAC condition** | | | | | | | | | | | | | | | **Z test** | |
|  | | | | | β | | | SE | | | t-stat | | | p*_FDR_* | | | β | | | | | | SE | | | t-stat | | | p*_FDR_* | | | p | |
| **Reappraisal** | | | | | .004 | | | .012 | | | .348 | | | .925 | | | -.006 | | | | | | .012 | | | -.496 | | | .803 | | | .552 | |
| **TIV** | | | | | .001 | | | <.001 | | | 3.679 | | | .008* | | | .001 | | | | | | <.001 | | | 3.674 | | | .004* | | | .964 | |
| **Age** | | | | | .002 | | | .011 | | | .134 | | | .925 | | | -.005 | | | | | | .011 | | | -.433 | | | .803 | | | .690 | |
| **Model** | | | | | N = 27, R² = .384, F = 4.7,  p = .010* | | | | | | | | | | | | | | | N = 27, R² = .400, F = 5.1,  p = .008* | | | | | | | | | | | | | |
| **Left striatum** \| \| \| \| \| \| \| \| \| \| \|; \| **E2 condition** \| \| \| \| \| **PLAC condition** \| \| \| \| \| **Z test**; \| \| β \| SE \| t-stat \| p*_FDR_* \| β \| \| SE \| t-stat \| p*_FDR_* \| p; **Reappraisal** \| \| -.102 \| .039 \| -2.600 \| .032 \| -.081 \| \| .036 \| -2.269 \| .049 \| .696; **TIV** \| \| .006 \| .001 \| 6.136 \| <.001* \| .066 \| \| .001 \| 6.7224 \| <.001* \| .999; **Age** \| \| .031 \| .045 \| .889 \| .383 \| .035 \| \| .032 \| 1.669 \| .3552 \| .949; **Model** \| \| N = 27, R² = .653, F = 14.4, p < .001* \| \| \| \| \| N = 27, R² = .681, F= 16.4, P < .001* \| \| \| \| | | | | | | | | | | | | | | | | | | | | | | | | | | | | | | | | | |
| **Right striatum** \| \| \| \| \| \| \| \| \| \| \|; \| **E2 condition** \| \| \| \| \| **PLAC condition** \| \| \| \| \| **Z test**; \| \| β \| SE \| t-stat \| p_FDR_ \| β \| \| SE \| t-stat \| p*_FDR_* \| p; **Reappraisal** \| \| -.114 \| .038 \| -3.029 \| .012* \| -.090 \| \| .039 \| -2.353 \| .041 \| .650; **TIV** \| \| .006 \| .001 \| 6.557 \| <.001* \| .007 \| \| .001 \| 6.743 \| <.001 \| .847; **Age** \| \| .025 \| .034 \| .744 \| .557 \| .015 \| \| .035 \| .441 \| .663 \| .845; **Model** \| \| N = 27, R² = .689, F = 17, p < .001* \| \| \| \| \| N = 27, R² = .689, F= 17, p < .001* \| \| \| \| | | | | | | | | | | | | | | | | | | | | | | | | | | | | | | | | | |
| **Left ventral striatum** | | | | | | | | | | | | | | | | | | | | | | | | | | | | | | | | | |
|  | **E2 condition** | | | | | | | | | | | | | | | **PLAC condition** | | | | | | | | | | | | | | | **Z test** | | |
|  | | | | β | | | SE | | | t-stat | | | p*_FDR_* | | | β | | | | | | SE | | | t-stat | | | p*_FDR_* | | | p | | |
| **Reappraisal** | | | | -.004 | | | .007 | | | -.565 | | | .694 | | | -.007 | | | | | | .006 | | | -1.114 | | | .415 | | | .733 | | |
| **TIV** | | | | .001 | | | <.001 | | | 4.262 | | | .001* | | | <.001 | | | | | | < .001 | | | 4.516 | | | .001* | | | .947 | | |
| **Age** | | | | .001 | | | .006 | | | .355 | | | .726 | | | -.001 | | | | | | .006 | | | -.260 | | | .797 | | | .662 | | |
| **Model** | | | | N = 27, R² = .448, F = 6.2,  p = .003* | | | | | | | | | | | | | | | N = 27, R² = .499, F = 7.6,  p = .001* | | | | | | | | | | | | | | |
| **Right ventral striatum** | | | | | | | | | | | | | | | | | | | | | | | | | | | | | | | | | |
|  | **E2 condition** | | | | | | | | | | | | | | | **PLAC condition** | | | | | | | | | | | | | | | **Z test** | | |
|  | | | | β | | | SE | | | t-stat | | | p*_FDR_* | | | β | | | | | | SE | | | t-stat | | | p*_FDR_* | | | p | | |
| **Reappraisal** | | | | -.012 | | | .005 | | | -2.400 | | | .050* | | | -.014 | | | | | | .006 | | | -2.204 | | | .076 | | | .836 | | |
| **TIV** | | | | .001 | | | <.001 | | | 5.804 | | | <.001* | | | .001 | | | | | | <.001 | | | 4.435 | | | .001* | | | .847 | | |
| **Age** | | | | .003 | | | .005 | | | .585 | | | .694 | | | .002 | | | | | | .006 | | | -.294 | | | .797 | | | .551 | | |
| **Model** | | | | N = 27, R² = .628, F = 12.9,  p < .001* | | | | | | | | | | | | | | | N = 27, R² = .526, F = 8.5,  p < .001* | | | | | | | | | | | | | | |
| **Left dorsal striatum** | | | | | | | | | | | | | | | | | | | | | | | | | | | | | | | | | |
|  | **E2 condition** | | | | | | | | | | | | | | | **PLAC condition** | | | | | | | | | | | | | | | **Z test** | | |
|  | | | | β | | | SE | | | t-stat | | | p*_FDR_* | | | β | | | | | | SE | | | t-stat | | | p*_FDR_* | | | p | | |
| **Reappraisal** | | | | -.092 | | | .035 | | | -2.649 | | | .024* | | | -.070 | | | | | | .032 | | | -2.223 | | | .060 | | | .651 | | |
| **TIV** | | | | .005 | | | .001 | | | 5.747 | | | <.001* | | | .005 | | | | | | > .001 | | | 6.320 | | | <.001* | | | .976 | | |
| **Age** | | | | .024 | | | .031 | | | .775 | | | .535 | | | .035 | | | | | | .029 | | | 1.237 | | | .274 | | | .792 | | |
| **Model** | | | | N = 27, R² = .630, F = 13.0,  p < .001* | | | | | | | | | | | | | | | N = 27, R² = .655, F = 14.6,  p < .001* | | | | | | | | | | | | | | |
| **Right dorsal striatum** | | | | | | | | | | | | | | | | | | | | | | | | | | | | | | | | | |
|  | **E2 condition** | | | | | | | | | | | | | | | **PLAC condition** | | | | | | | | | | | | | | | **Z test** | | |
|  | | | | β | | | SE | | | t-stat | | | p*_FDR_* | | | β | | | | | | SE | | | t-stat | | | p*_FDR_* | | | p | | |
| **Reappraisal** | | | | -.085 | | | .033 | | | -2.607 | | | .024* | | | -.071 | | | | | | .0323 | | | -2.177 | | | .060 | | | .766 | | |
| **TIV** | | | | .005 | | | .001 | | | 6.248 | | | <.001* | | | .008 | | | | | | .001 | | | 6.597 | | | <.001* | | | .791 | | |
| **Age** | | | | .005 | | | .030 | | | .174 | | | .863 | | | .013 | | | | | | .030 | | | .438 | | | .665 | | | .851 | | |
| **Model** | | | | N = 27, R² = .667, F = 15.4,  p < .001* | | | | | | | | | | | | | | | N = 27, R² = .678, F = 16.1,  p < .001* | | | | | | | | | | | | | | |
| **Left caudoventral striatum** | | | | | | | | | | | | | | | | | | | | | | | | | | | | | | | | | |
|  | **E2 condition** | | | | | | | | | | | | | | | **PLAC condition** | | | | | | | | | | | | | | | **Z test** | | |
|  | | | | β | | | SE | | | t-stat | | | p*_FDR_* | | | β | | | | | | SE | | | t-stat | | | p*_FDR_* | | | p | | |
| **Reappraisal** | | | | -.005 | | | .002 | | | -2.731 | | | .018* | | | -.004 | | | | | | .002 | | | -2.104 | | | .093 | | | .636 | | |
| **TIV** | | | | <.001 | | | <.001 | | | 5.914 | | | <.001* | | | <.001 | | | | | | <.001 | | | 5.605 | | | >.001* | | | .772 | | |
| **Age** | | | | .002 | | | .002 | | | 1.291 | | | .252 | | | <.001 | | | | | | .002 | | | -.139 | | | .890 | | | .309 | | |
| **Model** | | | | N = 27, R² = .643, F = 13.8,  p < .001* | | | | | | | | | | | | | | | N = 27, R² = .616, F = 12.3,  p < .001* | | | | | | | | | | | | | | |
| **Right caudoventral striatum** | | | | | | | | | | | | | | | | | | | | | | | | | | | | | | | | | |
|  | **E2 condition** | | | | | | | | | | | | | | | **PLAC condition** | | | | | | | | | | | | | | | **Z test** | | |
|  | | | | β | | | SE | | | t-stat | | | p*_FDR_* | | | β | | | | | | SE | | | t-stat | | | p*_FDR_* | | | p | | |
| **Reappraisal** | | | | -.006 | | | .002 | | | 3-382 | | | .005* | | | -.003 | | | | | | .002 | | | -1.121 | | | .411 | | | .207 | | |
| **TIV** | | | | <.001 | | | <.001 | | | 5.041 | | | <.001* | | | <.001 | | | | | | <.001 | | | 4.921 | | | <.001* | | | .526 | | |
| **Age** | | | | .001 | | | .002 | | | .866 | | | .396 | | | -.001 | | | | | | .002 | | | -.485 | | | .759 | | | .356 | | |
| **Model** | | | | N = 27, R² = .606, F = 11.8,  p < .001* | | | | | | | | | | | | | | | N = 27, R² = .547, F = 9.2,  p < .001* | | | | | | | | | | | | | | |

### Rumination

**Tab. S5 | Statistical parameters from robust mixed linear regression analysing** the relationship between emotion regulation strategy (trait reappraisal and rumination) and regional gray matter volume (GMV) under E2 and placebo (PLAC) conditions. Slope coefficients (β), standard errors (SE), t-statistics, and false discovery rate (FDR)-corrected p-values for the association between E2 increase and GMV across both drug conditions are reported. Additionally, p-values from Z-transformed slope comparisons (E2 vs. PLAC) are included to assess condition-specific differences in association strength. Significant associations (p < .05) are marked with an asterisk (*).

| **Left amygdala** | | | | | | | | | | | | | | | | | | | | | | | | | | | | | | | | | |
| --- | --- | --- | --- | --- | --- | --- | --- | --- | --- | --- | --- | --- | --- | --- | --- | --- | --- | --- | --- | --- | --- | --- | --- | --- | --- | --- | --- | --- | --- | --- | --- | --- | --- |
|  | | | **E2 condition** | | | | | | | | | | | | | | | **PLAC condition** | | | | | | | | | | | | | | | **Z test** |
|  | | | | | | β | | | SE | | | t-stat | | | p*_FDR_* | | | β | | | | | | SE | | | t-stat | | | p*_FDR_* | | | p |
| **Rumination** | | | | | | .007 | | | .007 | | | 1.028 | | | .472 | | | .006 | | | | | | .007 | | | .786 | | | .642 | | | .912 |
| **TIV** | | | | | | .001 | | | <.001 | | | 5.593 | | | <.001* | | | .001 | | | | | | <.001 | | | 4.903 | | | .002* | | | .892 |
| **Age** | | | | | | .005 | | | .008 | | | .668 | | | .511 | | | .006 | | | | | | .009 | | | .639 | | | .642 | | | .981 |
| **Model** | | | | | | N = 27, R² = .586, F = 10.8,  p < .001* | | | | | | | | | | | | | | | N = 27, R² = .518, F= 8.2,  p < .001* | | | | | | | | | | | | |
| **Right amygdala** | | | | | | | | | | | | | | | | | | | | | | | | | | | | | | | | | |
|  | | | **E2 condition** | | | | | | | | | | | | | | | **PLAC condition** | | | | | | | | | | | | | | | **Z test** |
|  | | | | | | β | | | SE | | | t-stat | | | p*_FDR_* | | | β | | | | | | SE | | | t-stat | | | p*_FDR_* | | | p |
| **Rumination** | | | | | | .006 | | | .008 | | | .775 | | | .511 | | | .004 | | | | | | .009 | | | .472 | | | .642 | | | .833 |
| **TIV** | | | | | | .002 | | | <.001 | | | 5.534 | | | <.001* | | | .002 | | | | | | <.001 | | | 5.230 | | | <.001* | | | .857 |
| **Age** | | | | | | .011 | | | .010 | | | 1.111 | | | .472 | | | .007 | | | | | | .010 | | | .583 | | | .642 | | | .713 |
| **Model** | | | | | | N = 27, R² = .575, F = 10.4,  p < .001* | | | | | | | | | | | | | | | N = 27, R² = .548, F = 9.3,  p < .001* | | | | | | | | | | | | |
| **Left ACC** \| \| \| \| \| \| \| \| \| \| \|; \| **E2 condition** \| \| \| \| \| **PLAC condition** \| \| \| \| \| **Z test**; \| \| β \| SE \| t-stat \| p*_FDR_* \| β \| \| SE \| t-stat \| p*_FDR_* \| p; **Rumination** \| \| .017 \| .030 \| .568 \| .619 \| .014 \| \| .029 \| .463 \| .747 \| .615; **TIV** \| \| .006 \| .001 \| 5.585 \| <.001 \| .006 \| \| .001 \| 5.784 \| <.001 \| .899; **Age** \| \| -.051 \| .035 \| -1.457 \| .317 \| -.059 \| \| .035 \| -1.699 \| .206 \| .914; **Model** \| \| N = 27, R² = .631, F = 13.1, p < .001* \| \| \| \| \| N = 27, R² = .652, F= 14.4, p < .001* \| \| \| \| | | | | | | | | | | | | | | | | | | | | | | | | | | | | | | | | | |
| **Right ACC** \| \| \| \| \| \| \| \| \| \| \|; \| **E2 condition** \| \| \| \| \| **PLAC condition** \| \| \| \| \| **Z test**; \| \| β \| SE \| t-stat \| p*_FDR_* \| β \| \| SE \| t-stat \| p*_FDR_* \| p; **Rumination** \| \| .014 \| .028 \| .504 \| .619 \| .009 \| \| .027 \| .327 \| .747 \| .791; **TIV** \| \| .006 \| .001 \| 5.846 \| <.001 \| .006 \| \| .001 \| 6.430 \| <.001 \| .902; **Age** \| \| -.018 \| .033 \| -.536 \| .619 \| -.025 \| \| .031 \| -.789 \| .658 \| .935; **Model** \| \| N = 27, R² = .623, F = 12.7, p < .001* \| \| \| \| \| N = 27, R² = .67, F = 15.6, p < .001* \| \| \| \| | | | | | | | | | | | | | | | | | | | | | | | | | | | | | | | | | |
| **Left perigenual ACC** | | | | | | | | | | | | | | | | | | | | | | | | | | | | | | | | | |
|  | | **E2 condition** | | | | | | | | | | | | | | | **PLAC condition** | | | | | | | | | | | | | | | **Z test** | |
|  | | | | | β | | | SE | | | t-stat | | | p*_FDR_* | | | β | | | | | | SE | | | t-stat | | | p*_FDR_* | | | p | |
| **Rumination** | | | | | .005 | | | .021 | | | .232 | | | .819 | | | .003 | | | | | | .021 | | | .146 | | | .959 | | | .954 | |
| **TIV** | | | | | .004 | | | .001 | | | 5.536 | | | <.001* | | | .004 | | | | | | .001 | | | 5.550 | | | <.001* | | | .916 | |
| **Age** | | | | | -.042 | | | .024 | | | -1.752 | | | .186 | | | -.047 | | | | | | .025 | | | -1.873 | | | .148 | | | .907 | |
| **Model** | | | | | N = 27, R² = .636, F = 13.4,  p < .001* | | | | | | | | | | | | | | | N = 27, R² = .642, F = 13.7,  p < .001* | | | | | | | | | | | | | |
| **Right perigenual ACC** | | | | | | | | | | | | | | | | | | | | | | | | | | | | | | | | | |
|  | | **E2 condition** | | | | | | | | | | | | | | | **PLAC condition** | | | | | | | | | | | | | | | **Z test** | |
|  | | | | | β | | | SE | | | t-stat | | | p*_FDR_* | | | β | | | | | | SE | | | t-stat | | | *p_FDR_* | | | p | |
| **Rumination** | | | | | .005 | | | .020 | | | .265 | | | .819 | | | .001 | | | | | | .019 | | | .052 | | | .959 | | | .878 | |
| **TIV** | | | | | .004 | | | .001 | | | 5.64 | | | <.001* | | | .004 | | | | | | .001 | | | 6.072 | | | <.001* | | | .829 | |
| **Age** | | | | | -.016 | | | .023 | | | -.698 | | | .738 | | | -.021 | | | | | | .023 | | | -.940 | | | .535 | | | .874 | |
| **Model** | | | | | N = 27, R² = .610, F = 12.0,  p < .001* | | | | | | | | | | | | | | | N = 27, R² = .650, F = 14.2,  p < .001* | | | | | | | | | | | | | |
| **Left subgenual ACC** | | | | | | | | | | | | | | | | | | | | | | | | | | | | | | | | | |
|  | | **E2 condition** | | | | | | | | | | | | | | | **PLAC condition** | | | | | | | | | | | | | | | **Z test** | |
|  | | | | | β | | | SE | | | t-stat | | | p*_FDR_* | | | β | | | | | | SE | | | t-stat | | | p*_FDR_* | | | p | |
| **Rumination** | | | | | .012 | | | .009 | | | 1.273 | | | .431 | | | .011 | | | | | | .009 | | | 1.223 | | | .340 | | | .947 | |
| **TIV** | | | | | .001 | | | <.001 | | | 4.386 | | | .001* | | | .001 | | | | | | <.001 | | | 4.622 | | | <.001* | | | .953 | |
| **Age** | | | | | -.004 | | | .011 | | | -.383 | | | .706 | | | -.009 | | | | | | .011 | | | -.858 | | | .340 | | | .748 | |
| **Model** | | | | | N = 27, R² = .623, F = 10.6,  p < .001* | | | | | | | | | | | | | | | N = 27, R² = .546, F = 8.8,  p < .001* | | | | | | | | | | | | | |
| **Right subgenual ACC** | | | | | | | | | | | | | | | | | | | | | | | | | | | | | | | | | |
|  | | **E2 condition** | | | | | | | | | | | | | | | **PLAC condition** | | | | | | | | | | | | | | | **Z test** | |
|  | | | | | β | | | SE | | | t-stat | | | p*_FDR_* | | | β | | | | | | SE | | | t-stat | | | p*_FDR_* | | | p | |
| **Rumination** | | | | | .007 | | | .007 | | | .968 | | | .515 | | | .006 | | | | | | .007 | | | .951 | | | .400 | | | .973 | |
| **TIV** | | | | | .001 | | | <.001 | | | 5.618 | | | <.001* | | | .001 | | | | | | <.001 | | | 5.877 | | | <.001* | | | .956 | |
| **Age** | | | | | -.004 | | | .008 | | | -.523 | | | .706 | | | -.008 | | | | | | .008 | | | -1.006 | | | .400 | | | .746 | |
| **Model** | | | | | N = 27, R² = .609, F = 16.9,  p < .001* | | | | | | | | | | | | | | | N = 27, R² = .641, F = 13.7,  p < .001* | | | | | | | | | | | | | |
| **Left dorsal ACC** | | | | | | | | | | | | | | | | | | | | | | | | | | | | | | | | | |
|  | | **E2 condition** | | | | | | | | | | | | | | | **PLAC condition** | | | | | | | | | | | | | | | **Z test** | |
|  | | | | | β | | | SE | | | t-stat | | | p*_FDR_* | | | β | | | | | | SE | | | t-stat | | | p*_FDR_* | | | p | |
| **Rumination** | | | | | <.001 | | | .003 | | | .074 | | | .942 | | | -.001 | | | | | | .003 | | | -.244 | | | .809 | | | .828 | |
| **TIV** | | | | | <.001 | | | <.001 | | | 3.923 | | | .004* | | | <.001 | | | | | | <.001 | | | 4.514 | | | .001* | | | .931 | |
| **Age** | | | | | -.004 | | | .004 | | | -1.194 | | | .490 | | | -.005 | | | | | | .003 | | | -1.361 | | | .373 | | | .986 | |
| **Model** | | | | | N = 27, R² = .497, F = 7.6,  p = .001* | | | | | | | | | | | | | | | N = 27, R² = .535, F = 8.8,  p < .001* | | | | | | | | | | | | | |
| **Right dorsal ACC** | | | | | | | | | | | | | | | | | | | | | | | | | | | | | | | | | |
|  | | **E2 condition** | | | | | | | | | | | | | | | **PLAC condition** | | | | | | | | | | | | | | | **Z test** | |
|  | | | | | β | | | SE | | | t-stat | | | p*_FDR_* | | | β | | | | | | SE | | | t-stat | | | p*_FDR_* | | | p | |
| **Rumination** | | | | | .002 | | | .004 | | | .423 | | | .811 | | | .001 | | | | | | .004 | | | .266 | | | .809 | | | .912 | |
| **TIV** | | | | | <.001 | | | <.001 | | | 3.609 | | | .004* | | | .001 | | | | | | <.001 | | | 3.627 | | | .004* | | | .981 | |
| **Age** | | | | | .002 | | | .005 | | | .493 | | | .811 | | | .003 | | | | | | .005 | | | .574 | | | .809 | | | .953 | |
| **Model** | | | | | N = 27, R² = .366, F = 4.4,  p = .013* | | | | | | | | | | | | | | | N = 27, R² =.367, F = 4.4,  p =.013* | | | | | | | | | | | | | |
| **Left total hippocampus** \| \| \| \| \| \| \| \| \| \| \|; \| **E2 condition** \| \| \| \| \| **PLAC condition** \| \| \| \| \| **Z test**; \| \| β \| SE \| t-stat \| p*_FDR_* \| β \| \| SE \| t-stat \| p*_FDR_* \| p; **Rumination** \| \| .030 \| .018 \| 1.644 \| .228 \| .025 \| \| .016 \| 1.530 \| .274 \| .897; **TIV** \| \| .003 \| .001 \| 4.513 \| <.001 \| .003 \| \| .001 \| 4.867 \| <.001 \| .997; **Age** \| \| .020 \| .021 \| .932 \| .541 \| .015 \| \| .019 \| .762 \| .545 \| .898; **Model** \| \| N = 27, R² = .498, F = 7.6, p = .001* \| \| \| \| \| N = 27, R² = .653, F= 8.6, p = .001* \| \| \| \| | | | | | | | | | | | | | | | | | | | | | | | | | | | | | | | | | |
| **Right total hippocampus** \| \| \| \| \| \| \| \| \| \| \|; \| **E2 condition** \| \| \| \| \| **PLAC condition** \| \| \| \| \| **Z test**; \| \| β \| SE \| t-stat \| p*_FDR_* \| β \| \| SE \| t-stat \| p*_FDR_* \| p; **Rumination** \| \| .013 \| .018 \| .752 \| .552 \| .019 \| \| .016 \| 1.150 \| .393 \| .720; **TIV** \| \| .003 \| .001 \| 4.498 \| <.001 \| .003 \| \| .001 \| 4.507 \| <.001 \| .833; **Age** \| \| .006 \| .021 \| .293 \| .772 \| -.003 \| \| .019 \| -.148 \| .884 \| .765; **Model** \| \| N = 27, R² = .481, F = 7.1, p = .001* \| \| \| \| \| N = 27, R² = .501, F= 7.7, p = .001* \| \| \| \| | | | | | | | | | | | | | | | | | | | | | | | | | | | | | | | | | |
| **Left hippocampus** | | | | | | | | | | | | | | | | | | | | | | | | | | | | | | | | | |
|  | | **E2 condition** | | | | | | | | | | | | | | | **PLAC condition** | | | | | | | | | | | | | | | **Z test** | |
|  | | | | | β | | | SE | | | t-stat | | | p*_FDR_* | | | β | | | | | | SE | | | t-stat | | | p*_FDR_* | | | p | |
| **Rumination** | | | | | .008 | | | .010 | | | .836 | | | .567 | | | .007 | | | | | | .010 | | | .670 | | | .677 | | | .905 | |
| **TIV** | | | | | .002 | | | <.001 | | | 4.820 | | | <.001* | | | .002 | | | | | | <.001 | | | 4.434 | | | .001* | | | .770 | |
| **Age** | | | | | .010 | | | .012 | | | .816 | | | .567 | | | .007 | | | | | | .012 | | | .587 | | | .677 | | | .869 | |
| **Model** | | | | | N = 27, R² = .465, F = 6.7,  p = .002* | | | | | | | | | | | | | | | N = 27, R² = .466, F = 6.7,  p = .002* | | | | | | | | | | | | | |
| **Right hippocampus** | | | | | | | | | | | | | | | | | | | | | | | | | | | | | | | | | |
|  | | **E2 condition** | | | | | | | | | | | | | | | **PLAC condition** | | | | | | | | | | | | | | | **Z test** | |
|  | | | | | β | | | SE | | | t-stat | | | p*_FDR_* | | | β | | | | | | SE | | | t-stat | | | p*_FDR_* | | | p | |
| **Rumination** | | | | | .008 | | | .010 | | | .731 | | | .567 | | | .007 | | | | | | .010 | | | .726 | | | .677 | | | .615 | |
| **TIV** | | | | | .002 | | | <.001 | | | 4.575 | | | <.001* | | | .001 | | | | | | <.001 | | | 4.093 | | | .001* | | | .899 | |
| **Age** | | | | | .007 | | | .012 | | | .566 | | | .577 | | | .003 | | | | | | .012 | | | .264 | | | .794 | | | .914 | |
| **Model** | | | | | N = 27, R² = .484, F = 7.2,  p = .001* | | | | | | | | | | | | | | | N = 27, R² = .436, F = 5.9,  p = .004* | | | | | | | | | | | | | |
| **Left para-hippocampus** | | | | | | | | | | | | | | | | | | | | | | | | | | | | | | | | | |
|  | | **E2 condition** | | | | | | | | | | | | | | | **PLAC condition** | | | | | | | | | | | | | | | **Z test** | |
|  | | | | | β | | | SE | | | t-stat | | | p*_FDR_* | | | β | | | | | | SE | | | t-stat | | | p*_FDR_* | | | p | |
| **Rumination** | | | | | .022 | | | .010 | | | 2.199 | | | .076 | | | .018 | | | | | | .009 | | | 2.082 | | | .097 | | | .792 | |
| **TIV** | | | | | .001 | | | <.001 | | | 3.599 | | | .005* | | | .001 | | | | | | <.001 | | | 4.228 | | | .002* | | | .909 | |
| **Age** | | | | | .013 | | | .012 | | | 1.070 | | | .444 | | | .009 | | | | | | .010 | | | .898 | | | .097 | | | .838 | |
| **Model** | | | | | N = 27, R² = .509, F = 8.0  p < .001* | | | | | | | | | | | | | | | N = 27, R² = .487, F = 7.3,  p = .001* | | | | | | | | | | | | | |
| **Right para-hippocampus** | | | | | | | | | | | | | | | | | | | | | | | | | | | | | | | | | |
|  | | **E2 condition** | | | | | | | | | | | | | | | **PLAC condition** | | | | | | | | | | | | | | | **Z test** | |
|  | | | | | β | | | SE | | | t-stat | | | p*_FDR_* | | | β | | | | | | SE | | | t-stat | | | p*_FDR_* | | | p | |
| **Rumination** | | | | | .006 | | | .009 | | | .624 | | | .646 | | | .012 | | | | | | .009 | | | 1.352 | | | .284 | | | .629 | |
| **TIV** | | | | | .001 | | | <.001 | | | 3.733 | | | .004* | | | .001 | | | | | | <.001 | | | 3.716 | | | .003* | | | .898 | |
| **Age** | | | | | .002 | | | .022 | | | .183 | | | .856 | | | -.005 | | | | | | .011 | | | -.444 | | | .662 | | | .661 | |
| **Model** | | | | | N = 27, R² = .390, F = 4.9,  p = .009* | | | | | | | | | | | | | | | N = 27, R² = .428, F = 5.7,  p = .004* | | | | | | | | | | | | | |
| (**Left striatum** \| \| \| \| \| \| \| \| \| \| \|; \| **E2 condition** \| \| \| \| \| **PLAC condition** \| \| \| \| \| **Z test**; \| \| β \| SE \| t-stat \| p*_FDR_* \| β \| \| SE \| t-stat \| p*_FDR_* \| p; **Rumination** \| \| .030 \| .032 \| .953 \| .526 \| .019 \| \| .028 \| .695 \| .750 \| .791; **TIV** \| \| .006 \| .001 \| 5.763 \| <.001* \| .006 \| \| .001 \| 6.597 \| <.001* \| .987; **Age** \| \| .013 \| .038 \| .344 \| .734 \| .011 \| \| .033 \| .341 \| .795 \| .973; **Model** \| \| N = 27, R² = .604, F = 11.7, p < .001* \| \| \| \| \| N = 27, R² = .664, F = 15.1, p < .001* \| \| \| \|) | | | | | | | | | | | | | | | | | | | | | | | | | | | | | | | | | |
| **Right striatum** \| \| \| \| \| \| \| \| \| \| \|; \| **E2 condition** \| \| \| \| \| **PLAC condition** \| \| \| \| \| **Z test**; \| \| β \| SE \| t-stat \| p*_FDR_* \| β \| \| SE \| t-stat \| p*_FDR_* \| p; **Rumination** \| \| .052 \| .029 \| 1.811 \| .166 \| .022 \| \| .031 \| .717 \| .750 \| .491; **TIV** \| \| .007 \| .001 \| 7.277 \| <.001* \| .007 \| \| .001 \| 6.453 \| <.001* \| .900; **Age** \| \| .024 \| .034 \| .702 \| .588 \| .010 \| \| .037 \| .262 \| .795 \| .781; **Model** \| \| N = 27, R² = .714, F = 19.1, p < .001* \| \| \| \| \| N = 27, R² = .656, F = 14.6, p < .001* \| \| \| \| | | | | | | | | | | | | | | | | | | | | | | | | | | | | | | | | | |
| **Left ventral striatum** | | | | | | | | | | | | | | | | | | | | | | | | | | | | | | | | | |
|  | **E2 condition** | | | | | | | | | | | | | | | **PLAC condition** | | | | | | | | | | | | | | | **Z test** | | |
|  | | | | β | | | SE | | | t-stat | | | p*_FDR_* | | | β | | | | | | SE | | | t-stat | | | p*_FDR_* | | | p | | |
| **Rumination** | | | | .005 | | | .005 | | | .947 | | | .426 | | | .004 | | | | | | .005 | | | .874 | | | .783 | | | .912 | | |
| **TIV** | | | | .001 | | | <.001 | | | 4.391 | | | .001* | | | .001 | | | | | | <.001 | | | 4.777 | | | .001* | | | .981 | | |
| **Age** | | | | .002 | | | .006 | | | .319 | | | .753 | | | -.002 | | | | | | .006 | | | -.371 | | | .857 | | | .627 | | |
| **Model** | | | | N = 27, R² = .472, F = 6.8,  p = .002* | | | | | | | | | | | | | | | N = 27, R² = .528, F = 8.6,  p < .001* | | | | | | | | | | | | | | |
| **Right ventral striatum** | | | | | | | | | | | | | | | | | | | | | | | | | | | | | | | | | |
|  | **E2 condition** | | | | | | | | | | | | | | | **PLAC condition** | | | | | | | | | | | | | | | **Z test** | | |
|  | | | | β | | | SE | | | t-stat | | | p*_FDR_* | | | β | | | | | | SE | | | t-stat | | | p*_FDR_* | | | p | | |
| **Rumination** | | | | .005 | | | .004 | | | 1.068 | | | .426 | | | -.001 | | | | | | .006 | | | -.097 | | | .924 | | | .466 | | |
| **TIV** | | | | .001 | | | <.001 | | | 5.903 | | | <.001* | | | .001 | | | | | | <.001 | | | 3.740 | | | .003* | | | .515 | | |
| **Age** | | | | .005 | | | .005 | | | .944 | | | .426 | | | -.003 | | | | | | .007 | | | -.527 | | | .857 | | | .320 | | |
| **Model** | | | | N = 27, R² = .613, F = 12.1,  p < .001* | | | | | | | | | | | | | | | N = 27, R² = .411, F = 5.3,  p = .006* | | | | | | | | | | | | | | |
| **Left dorsal striatum** | | | | | | | | | | | | | | | | | | | | | | | | | | | | | | | | | |
|  | **E2 condition** | | | | | | | | | | | | | | | **PLAC condition** | | | | | | | | | | | | | | | **Z test** | | |
|  | | | | β | | | SE | | | t-stat | | | p*_FDR_* | | | β | | | | | | SE | | | t-stat | | | p*_FDR_* | | | p | | |
| **Rumination** | | | | .024 | | | .029 | | | .822 | | | .630 | | | .017 | | | | | | .025 | | | .685 | | | .723 | | | .856 | | |
| **TIV** | | | | .005 | | | .001 | | | 5.048 | | | <.001* | | | .005 | | | | | | .001 | | | 6.283 | | | <.001* | | | .820 | | |
| **Age** | | | | -.003 | | | .034 | | | -.098 | | | .923 | | | .015 | | | | | | .029 | | | .530 | | | .723 | | | .627 | | |
| **Model** | | | | N = 27, R² = .546, F = 9.2,  p < .001* | | | | | | | | | | | | | | | N = 27, R² = .640, F = 13.6,  p < .001* | | | | | | | | | | | | | | |
| **Right dorsal striatum** | | | | | | | | | | | | | | | | | | | | | | | | | | | | | | | | | |
|  | **E2 condition** | | | | | | | | | | | | | | | **PLAC condition** | | | | | | | | | | | | | | | **Z test** | | |
|  | | | | β | | | SE | | | t-stat | | | p*_FDR_* | | | β | | | | | | SE | | | t-stat | | | p*_FDR_* | | | p | | |
| **Rumination** | | | | .040 | | | .026 | | | 1.527 | | | .281 | | | .017 | | | | | | .027 | | | .648 | | | .722 | | | .541 | | |
| **TIV** | | | | .006 | | | .001 | | | 6.513 | | | <.001* | | | .006 | | | | | | .001 | | | 6.299 | | | <.001* | | | .930 | | |
| **Age** | | | | .012 | | | .031 | | | .381 | | | .848 | | | .003 | | | | | | .032 | | | .096 | | | .924 | | | .842 | | |
| **Model** | | | | N = 27 R² = .667, F = 15.3,  p < .001* | | | | | | | | | | | | | | | N = 27, R² = .647, F = 14.0,  p < .001* | | | | | | | | | | | | | | |
| **Left caudoventral striatum** | | | | | | | | | | | | | | | | | | | | | | | | | | | | | | | | | |
|  | **E2 condition** | | | | | | | | | | | | | | | **PLAC condition** | | | | | | | | | | | | | | | **Z test** | | |
|  | | | | β | | | SE | | | t-stat | | | p*_FDR_* | | | β | | | | | | SE | | | t-stat | | | p*_FDR_* | | | p | | |
| **Rumination** | | | | <.001 | | | .002 | | | .241 | | | .812 | | | .001 | | | | | | .002 | | | .494 | | | .715 | | | .857 | | |
| **TIV** | | | | <.001 | | | .001 | | | 5.066 | | | <.001* | | | <.001 | | | | | | <.001 | | | 4.812 | | | <.001* | | | .744 | | |
| **Age** | | | | .002 | | | .002 | | | 1.066 | | | .446 | | | -.001 | | | | | | .002 | | | -.370 | | | .715 | | | .305 | | |
| **Model** | | | | N = 27, R² = .534, F = 8.8,  p < .001* | | | | | | | | | | | | | | | N = 27, R² = .527, F = 8.5,  p < .001* | | | | | | | | | | | | | | |
| **Right caudoventral striatum** | | | | | | | | | | | | | | | | | | | | | | | | | | | | | | | | | |
|  | **E2 condition** | | | | | | | | | | | | | | | **PLAC condition** | | | | | | | | | | | | | | | **Z test** | | |
|  | | | | β | | | SE | | | t-stat | | | p*_FDR_* | | | β | | | | | | SE | | | t-stat | | | p*_FDR_* | | | p | | |
| **Rumination** | | | | .003 | | | .002 | | | 1.778 | | | .177 | | | .002 | | | | | | .002 | | | .940 | | | .714 | | | .574 | | |
| **TIV** | | | | <.001 | | | .001 | | | 3.997 | | | .002* | | | <.001 | | | | | | .001 | | | 4.738 | | | <.001* | | | .533 | | |
| **Age** | | | | .001 | | | .002 | | | .444 | | | .793 | | | -.001 | | | | | | .002 | | | -.610 | | | .715 | | | .455 | | |
| **Model** | | | | N = 27, R² = .454, F = 6.4,  p =.003* | | | | | | | | | | | | | | | N = 27, R² = .533, F = 8.7,  p < .001* | | | | | | | | | | | | | | |

Interaction E2 x reappraisal.

**Tab. S6 | Statistical parameters from robust mixed linear regression analysing** the relationship between emotion regulation strategy (trait reappraisal and rumination) by E2 increase and regional gray matter volume (GMV) under E2 and placebo (PLAC) conditions. Slope coefficients (β), standard errors (SE), t-statistics, and false discovery rate (FDR)-corrected p-values for the association between E2 increase and GMV across both drug conditions are reported. Additionally, p-values from Z-transformed slope comparisons (E2 vs. PLAC) are included to assess condition-specific differences in association strength. Significant associations (p < .05) are marked with an asterisk (*).

| **Left amygdala** | | | | | | | | | | | | | | | | | | | | | | | | | | | | | | | | | |
| --- | --- | --- | --- | --- | --- | --- | --- | --- | --- | --- | --- | --- | --- | --- | --- | --- | --- | --- | --- | --- | --- | --- | --- | --- | --- | --- | --- | --- | --- | --- | --- | --- | --- |
|  | | | **E2 condition** | | | | | | | | | | | | | | | **PLAC condition** | | | | | | | | | | | | | | | **Z test** |
|  | | | | | | β | | | SE | | | t-stat | | | p*_FDR_* | | | β | | | | | | SE | | | t-stat | | | p*_FDR_* | | | p |
| **Reappraisal x E2Δ** | | | | | | -.026 | | | .020 | | | -1.321 | | | .932 | | | -.004 | | | | | | -.009 | | | -.507 | | | .909 | | | .427 |
| **TIV** | | | | | | .002 | | | <.001 | | | 5.297 | | | <.001* | | | .001 | | | | | | <.001 | | | 4.921 | | | <.001* | | | .957 |
| **Age** | | | | | | .005 | | | .011 | | | .470 | | | .932 | | | .001 | | | | | | .011 | | | -.072 | | | .943 | | | .919 |
| **Model** | | | | | | N = 21, R² = .645, F = 10.3,  p < .001* | | | | | | | | | | | | | | | N = 24, R² = .558, F= 8.4,  p = .001* | | | | | | | | | | | | |
| **Right amygdala** | | | | | | | | | | | | | | | | | | | | | | | | | | | | | | | | | |
|  | | | **E2 condition** | | | | | | | | | | | | | | | **PLAC condition** | | | | | | | | | | | | | | | **Z test** |
|  | | | | | | β | | | SE | | | t-stat | | | p*_FDR_* | | | β | | | | | | SE | | | t-stat | | | p*_FDR_* | | | p |
| **Reappraisal x E2Δ** | | | | | | .002 | | | .023 | | | .092 | | | .932 | | | -.003 | | | | | | .009 | | | -.335 | | | 909 | | | .835 |
| **TIV** | | | | | | .002 | | | <.001 | | | 5.228 | | | <.001* | | | .002 | | | | | | <.001 | | | 5.545 | | | <.001* | | | .807 |
| **Age** | | | | | | -.002 | | | .023 | | | -.087 | | | .932 | | | -.004 | | | | | | .011 | | | -.313 | | | .909 | | | .880 |
| **Model** | | | | | | N = 21, R² = .640, F = 10.1,  p < .001* | | | | | | | | | | | | | | | N = 24, R² = .622, F = 11.0,  p < .001* | | | | | | | | | | | | |
| **Left ACC** \| \| \| \| \| \| \| \| \| \| \|; \| **E2 condition** \| \| \| \| \| **PLAC condition** \| \| \| \| \| **Z test**; \| \| β \| SE \| t-stat \| p*_FDR_* \| β \| \| SE \| t-stat \| p*_FDR_* \| p; **Reappraisal x E2Δ** \| \| -.084 \| .072 \| -1.176 \| .384 \| -.012 \| \| .030 \| -.402 \| .831 \| .351; **TIV** \| \| .007 \| .001 \| 6.785 \| <.001* \| .006 \| \| .001 \| 6.708 \| <.001* \| .706; **Age** \| \| -.087 \| .038 \| -2.308 \| .068 \| -.073 \| \| .038 \| -1.954 \| .130 \| .780; **Model** \| \| N = 21, R² = .784, F =20.5, p < .001* \| \| \| \| \| N = 24, R² = .736, F= 18.6, p < .001* \| \| \| \| | | | | | | | | | | | | | | | | | | | | | | | | | | | | | | | | | |
| **Right ACC** \| \| \| \| \| \| \| \| \| \| \|; \| **E2 condition** \| \| \| \| \| **PLAC condition** \| \| \| \| \| **Z test**; \| \| β \| SE \| t-stat \| p*_FDR_* \| β \| \| SE \| t-stat \| p*_FDR_* \| p; **Reappraisal x E2Δ** \| \| -.027 \| .075 \| -.362 \| .722 \| -.005 \| \| .025 \| -.213 \| .834 \| .783; **TIV** \| \| .006 \| .001 \| 6.107 \| <.001* \| .006 \| \| .001 \| 7.764 \| <.001* \| .882; **Age** \| \| -.038 \| .039 \| -.975 \| .412 \| -.034 \| \| .032 \| -1.06 \| .454 \| .930; **Model** \| \| N = 21, R² = .721, F = 14.6, p < .001* \| \| \| \| \| N = 24, R² = .772, F = 22.6, p < .001* \| \| \| \| | | | | | | | | | | | | | | | | | | | | | | | | | | | | | | | | | |
| **Left perigenual ACC** | | | | | | | | | | | | | | | | | | | | | | | | | | | | | | | | | |
|  | | **E2 condition** | | | | | | | | | | | | | | | **PLAC condition** | | | | | | | | | | | | | | | **Z test** | |
|  | | | | | β | | | SE | | | t-stat | | | p*_FDR_* | | | β | | | | | | SE | | | t-stat | | | p*_FDR_* | | | p | |
| **Reappraisal x E2Δ** | | | | | -.076 | | | .048 | | | -1.585 | | | .197 | | | -.016 | | | | | | .022 | | | -.711 | | | .583 | | | .256 | |
| **TIV** | | | | | .005 | | | .001 | | | 6.944 | | | <.001* | | | .004 | | | | | | .001 | | | 6.094 | | | <.001* | | | .704 | |
| **Age** | | | | | -.065 | | | .025 | | | -2.579 | | | .039* | | | -.058 | | | | | | .028 | | | -2.060 | | | .105 | | | .856 | |
| **Model** | | | | | N = 21, R² = .795, F = 4.4,  p < .001* | | | | | | | | | | | | | | | N = 24, R² = .703, F = 15.8,  p < .001* | | | | | | | | | | | | | |
| **Right perigenual ACC** | | | | | | | | | | | | | | | | | | | | | | | | | | | | | | | | | |
|  | | **E2 condition** | | | | | | | | | | | | | | | **PLAC condition** | | | | | | | | | | | | | | | **Z test** | |
|  | | | | | Β | | | SE | | | t-stat | | | p*_FDR_* | | | β | | | | | | SE | | | t-stat | | | *p_FDR_* | | | p | |
| **Reappraisal x E2Δ** | | | | | -.018 | | | .054 | | | -.328 | | | .747 | | | -.005 | | | | | | .019 | | | -.261 | | | .797 | | | .822 | |
| **TIV** | | | | | .004 | | | .001 | | | 5.567 | | | <.001* | | | .004 | | | | | | <.001 | | | 7.082 | | | <.001* | | | .953 | |
| **Age** | | | | | -.029 | | | .028 | | | -1.022 | | | .385 | | | -.028 | | | | | | .024 | | | -1.175 | | | .381 | | | .969 | |
| **Model** | | | | | N = 21, R² = .686, F =12.4,  p < .001* | | | | | | | | | | | | | | | N = 24, R² = .741, F = 19.1,  p < .001* | | | | | | | | | | | | | |
| **Left subgenual ACC** | | | | | | | | | | | | | | | | | | | | | | | | | | | | | | | | | |
|  | | **E2 condition** | | | | | | | | | | | | | | | **PLAC condition** | | | | | | | | | | | | | | | **Z test** | |
|  | | | | | β | | | SE | | | t-stat | | | p*_FDR_* | | | β | | | | | | SE | | | t-stat | | | p*_FDR_* | | | p | |
| **Reappraisal x E2Δ** | | | | | -.009 | | | .025 | | | -.363 | | | .866 | | | .004 | | | | | | .009 | | | .424 | | | .677 | | | .628 | |
| **TIV** | | | | | .002 | | | <.001 | | | 4.707 | | | .001* | | | .002 | | | | | | .001 | | | 5.197 | | | <.001* | | | .692 | |
| **Age** | | | | | -.015 | | | .013 | | | -1.109 | | | .566 | | | -.011 | | | | | | .012 | | | -.952 | | | .329 | | | .827 | |
| **Model** | | | | | N = 21, R² = .619, F = 9.2,  p = .001* | | | | | | | | | | | | | | | N = 24, R² = .621, F = 10.9,  p < .001* | | | | | | | | | | | | | |
| **Right subgenual ACC** | | | | | | | | | | | | | | | | | | | | | | | | | | | | | | | | | |
|  | | **E2 condition** | | | | | | | | | | | | | | | **PLAC condition** | | | | | | | | | | | | | | | **Z test** | |
|  | | | | | β | | | SE | | | t-stat | | | p*_FDR_* | | | β | | | | | | SE | | | t-stat | | | p*_FDR_* | | | p | |
| **Reappraisal x E2Δ** | | | | | -.001 | | | .020 | | | -.063 | | | .950 | | | .004 | | | | | | .007 | | | .498 | | | .677 | | | .827 | |
| **TIV** | | | | | .002 | | | <.001 | | | 5.664 | | | <.001* | | | .001 | | | | | | <.001 | | | 6.623 | | | <.001* | | | .608 | |
| **Age** | | | | | -.008 | | | .011 | | | -.789 | | | .661 | | | -.008 | | | | | | .009 | | | -.976 | | | .529 | | | 1 | |
| **Model** | | | | | N = 21, R² = .690, F = 12.6,  p < .001* | | | | | | | | | | | | | | | N = 24, R² = .721, F = 17.3,  p < .001* | | | | | | | | | | | | | |
| **Left dorsal ACC** | | | | | | | | | | | | | | | | | | | | | | | | | | | | | | | | | |
|  | | **E2 condition** | | | | | | | | | | | | | | | **PLAC condition** | | | | | | | | | | | | | | | **Z test** | |
|  | | | | | β | | | SE | | | t-stat | | | p*_FDR_* | | | β | | | | | | SE | | | t-stat | | | p*_FDR_* | | | p | |
| **Reappraisal x E2Δ** | | | | | .002 | | | .008 | | | .202 | | | .842 | | | .001 | | | | | | ,003 | | | .314 | | | .887 | | | .935 | |
| **TIV** | | | | | <.001 | | | <.001 | | | 4.252 | | | .002* | | | <.001 | | | | | | <.001 | | | 5.107 | | | <.001* | | | .990 | |
| **Age** | | | | | -.007 | | | .004 | | | -1.799 | | | .180 | | | -.005 | | | | | | .156 | | | -1.473 | | | .313 | | | .710 | |
| **Model** | | | | | N = 21, R² = .622, F = 9.3,  p = .001* | | | | | | | | | | | | | | | N = 24, R² = .628, F = 11.3,  p < .001* | | | | | | | | | | | | | |
| **Right dorsal ACC** | | | | | | | | | | | | | | | | | | | | | | | | | | | | | | | | | |
|  | | **E2 condition** | | | | | | | | | | | | | | | **PLAC condition** | | | | | | | | | | | | | | | **Z test** | |
|  | | | | | β | | | SE | | | t-stat | | | p*_FDR_* | | | β | | | | | | SE | | | t-stat | | | p*_FDR_* | | | p | |
| **Reappraisal x E2Δ** | | | | | -.003 | | | .010 | | | -.272 | | | .842 | | | -.003 | | | | | | .004 | | | -.639 | | | .759 | | | .995 | |
| **TIV** | | | | | .001 | | | <.001 | | | 4.093 | | | .002* | | | <.001 | | | | | | <.001 | | | 4.085 | | | .002* | | | .848 | |
| **Age** | | | | | -.002 | | | .005 | | | -.275 | | | .842 | | | <.001 | | | | | | .005 | | | .143 | | | .887 | | | .767 | |
| **Model** | | | | | N = 21, R² = .521, F = 6.2,  p = .005* | | | | | | | | | | | | | | | N = 24, R² =.460, F = 5.7,  p =.006* | | | | | | | | | | | | | |
| **Left total hippocampus** \| \| \| \| \| \| \| \| \| \| \|; \| **E2 condition** \| \| \| \| \| **PLAC condition** \| \| \| \| \| **Z test**; \| \| β \| SE \| t-stat \| p*_FDR_* \| β \| \| SE \| t-stat \| p*_FDR_* \| p; **Reappraisal x E2Δ** \| \| .007 \| .054 \| .128 \| .892 \| -.024 \| \| .019 \| -1.246 \| .455 \| .585; **TIV** \| \| .002 \| .001 \| 3.961 \| .003* \| .003 \| \| .001 \| 4.861 \| <.001* \| .957; **Age** \| \| -.007 \| .028 \| -.244 \| .892 \| -.003 \| \| .024 \| -.114 \| .910 \| .911; **Model** \| \| N = 21, R² = .513, F = 6.0, p = .006* \| \| \| \| \| N = 24, R² = .552, F= 8.2, p = .001* \| \| \| \| | | | | | | | | | | | | | | | | | | | | | | | | | | | | | | | | | |
| **Right total hippocampus** \| \| \| \| \| \| \| \| \| \| \|; \| **E2 condition** \| \| \| \| \| **PLAC condition** \| \| \| \| \| **Z test**; \| \| β \| SE \| t-stat \| p*_FDR_* \| β \| \| SE \| t-stat \| p*_FDR_* \| p; **Reappraisal x E2Δ** \| \| -.032 \| .055 \| -.580 \| .855 \| -.018 \| \| .019 \| -.946 \| .533 \| .816; **TIV** \| \| .003 \| .001 \| 4.074 \| <.001* \| .003 \| \| .001 \| 4.724 \| <.001* \| .795; **Age** \| \| -.017 \| .028 \| -.589 \| .855 \| -.012 \| \| .024 \| -.484 \| .760 \| .891; **Model** \| \| N = 21, R² = .528, F = 6.3, p = .004* \| \| \| \| \| N = 24, R² = .546, F= 8.0, p = .001* \| \| \| \| | | | | | | | | | | | | | | | | | | | | | | | | | | | | | | | | | |
| **Left hippocampus** | | | | | | | | | | | | | | | | | | | | | | | | | | | | | | | | | |
|  | | **E2 condition** | | | | | | | | | | | | | | | **PLAC condition** | | | | | | | | | | | | | | | **Z test** | |
|  | | | | | β | | | SE | | | t-stat | | | p*_FDR_* | | | β | | | | | | SE | | | t-stat | | | p*_FDR_* | | | p | |
| **Reappraisal x E2Δ** | | | | | .004 | | | .026 | | | .158 | | | .986 | | | -.009 | | | | | | .010 | | | -.821 | | | .633 | | | .654 | |
| **TIV** | | | | | .002 | | | <.001 | | | 4.956 | | | <.001* | | | .002 | | | | | | <.001 | | | 4.766 | | | <.001* | | | .588 | |
| **Age** | | | | | -.003 | | | .014 | | | -.244 | | | .986 | | | -.008 | | | | | | .013 | | | -.644 | | | .633 | | | .791 | |
| **Model** | | | | | N = 21, R² = .620, F = 9.3,  p = .001* | | | | | | | | | | | | | | | N = 24, R² = .554, F = 8.3,  p = .001* | | | | | | | | | | | | | |
| **Right hippocampus** | | | | | | | | | | | | | | | | | | | | | | | | | | | | | | | | | |
|  | | **E2 condition** | | | | | | | | | | | | | | | **PLAC condition** | | | | | | | | | | | | | | | **Z test** | |
|  | | | | | β | | | SE | | | t-stat | | | p*_FDR_* | | | β | | | | | | SE | | | t-stat | | | p*_FDR_* | | | p | |
| **Reappraisal x E2Δ** | | | | | -.014 | | | .032 | | | -.417 | | | .986 | | | -.009 | | | | | | .012 | | | -.737 | | | .633 | | | .900 | |
| **TIV** | | | | | .002 | | | <.001 | | | 4.126 | | | .002* | | | .002 | | | | | | <.001 | | | 4.077 | | | .002* | | | .648 | |
| **Age** | | | | | <-.001 | | | .017 | | | -.017 | | | .986 | | | <.001 | | | | | | .016 | | | -.003 | | | .998 | | | .991 | |
| **Model** | | | | | N = 21, R² = .515, F = 6.0,  p = .006* | | | | | | | | | | | | | | | N = 24, R² = .461, F = 5.7,  p = .006* | | | | | | | | | | | | | |
| **Left para-hippocampus** | | | | | | | | | | | | | | | | | | | | | | | | | | | | | | | | | |
|  | | **E2 condition** | | | | | | | | | | | | | | | **PLAC condition** | | | | | | | | | | | | | | | **Z test** | |
|  | | | | | β | | | SE | | | t-stat | | | p*_FDR_* | | | β | | | | | | SE | | | t-stat | | | p*_FDR_* | | | p | |
| **Reappraisal x E2Δ** | | | | | .003 | | | .032 | | | .086 | | | .933 | | | -.016 | | | | | | .010 | | | -1.618 | | | .243 | | | .575 | |
| **TIV** | | | | | .001 | | | <.001 | | | 2.601 | | | .056 | | | .001 | | | | | | <.001 | | | 4.510 | | | <.001* | | | .648 | |
| **Age** | | | | | -.003 | | | .017 | | | -.149 | | | .933 | | | .012 | | | | | | .013 | | | .842 | | | .482 | | | .533 | |
| **Model** | | | | | N = 21, R² = .312, F = 2.6,  p = 089 | | | | | | | | | | | | | | | N = 24, R² = .514, F = 7.0,  p = .002* | | | | | | | | | | | | | |
| **Right para-hippocampus** | | | | | | | | | | | | | | | | | | | | | | | | | | | | | | | | | |
|  | | **E2 condition** | | | | | | | | | | | | | | | **PLAC condition** | | | | | | | | | | | | | | | **Z test** | |
|  | | | | | β | | | SE | | | t-stat | | | p*_FDR_* | | | β | | | | | | SE | | | t-stat | | | p*_FDR_* | | | p | |
| **Reappraisal x E2Δ** | | | | | -.020 | | | .026 | | | -.784 | | | .666 | | | -.007 | | | | | | .009 | | | -1.002 | | | .478 | | | .700 | |
| **TIV** | | | | | .001 | | | <.001 | | | 3.528 | | | .016* | | | .001 | | | | | | <.001 | | | 4.505 | | | <.001* | | | .885 | |
| **Age** | | | | | -.015 | | | .014 | | | -1.137 | | | .543 | | | -.009 | | | | | | .012 | | | -.717 | | | .482 | | | .704 | |
| **Model** | | | | | N = 21, R² = .489, F = 5.4,  p = .008* | | | | | | | | | | | | | | | N = 24, R² = .529, F = 7.5,  p = .002* | | | | | | | | | | | | | |
| (**Left striatum** \| \| \| \| \| \| \| \| \| \| \|; \| **E2 condition** \| \| \| \| \| **PLAC condition** \| \| \| \| \| **Z test**; \| \| β \| SE \| t-stat \| p*_FDR_* \| β \| \| SE \| t-stat \| p*_FDR_* \| p; **Reappraisal x E2Δ** \| \| .001 \| .085 \| .011 \| .991 \| -009 \| \| .028 \| -.339 \| .855 \| .907; **TIV** \| \| .006 \| .001 \| 4.960 \| .001* \| .007 \| \| .001 \| 7.389 \| <.001* \| .677; **Age** \| \| -.013 \| .045 \| -.280 \| .940 \| -.011 \| \| .035 \| -.306 \| .855 \| .976; **Model** \| \| N = 21, R² = .622, F = 9.3, p =.001 * \| \| \| \| \| N = 24, R² = .747, F = 19.7, p < .001* \| \| \| \|) | | | | | | | | | | | | | | | | | | | | | | | | | | | | | | | | | |
| **Right striatum** \| \| \| \| \| \| \| \| \| \| \|; \| **E2 condition** \| \| \| \| \| **PLAC condition** \| \| \| \| \| **Z test**; \| \| β \| SE \| t-stat \| p*_FDR_* \| β \| \| SE \| t-stat \| p*_FDR_* \| p; **Reappraisal x E2Δ** \| \| .034 \| .094 \| .366 \| .940 \| .006 \| \| .031 \| .185 \| .855 \| .771; **TIV** \| \| .006 \| .001 \| 4.736 \| .001* \| .006 \| \| .001 \| 6.934 \| <.001* \| .759; **Age** \| \| -.027 \| .049 \| -.546 \| .940 \| -.037 \| \| .039 \| -.942 \| .715 \| .878; **Model** \| \| N = 21, R² = .615, F = 9.04, p = .001* \| \| \| \| \| N = 24, R² = .737., F = 18.7, p < .001* \| \| \| \| | | | | | | | | | | | | | | | | | | | | | | | | | | | | | | | | | |
| **Left ventral striatum** | | | | | | | | | | | | | | | | | | | | | | | | | | | | | | | | | |
|  | **E2 condition** | | | | | | | | | | | | | | | **PLAC condition** | | | | | | | | | | | | | | | **Z test** | | |
|  | | | | β | | | SE | | | t-stat | | | p*_FDR_* | | | β | | | | | | SE | | | t-stat | | | p*_FDR_* | | | p | | |
| **Reappraisal x E2Δ** | | | | .001 | | | .014 | | | .079 | | | .938 | | | .007 | | | | | | .005 | | | 1.424 | | | .219 | | | 6.88 | | |
| **TIV** | | | | .001* | | | <.001 | | | 2.879 | | | .031* | | | .001 | | | | | | <.001 | | | 4.951 | | | .001* | | | .399 | | |
| **Age** | | | | -.009 | | | .007 | | | -1.257 | | | .451 | | | -.005 | | | | | | .006 | | | -.758 | | | .457 | | | .643 | | |
| **Model** | | | | N = 21, R² = .431, F = 4.3,  p = .020* | | | | | | | | | | | | | | | N = 24, R² = .625, F = 11.1,  p < .001* | | | | | | | | | | | | | | |
| **Right ventral striatum** | | | | | | | | | | | | | | | | | | | | | | | | | | | | | | | | | |
|  | **E2 condition** | | | | | | | | | | | | | | | **PLAC condition** | | | | | | | | | | | | | | | **Z test** | | |
|  | | | | β | | | SE | | | t-stat | | | p*_FDR_* | | | β | | | | | | SE | | | t-stat | | | p*_FDR_* | | | p | | |
| **Reappraisal x E2Δ** | | | | -.003 | | | .013 | | | -.242 | | | .938 | | | .010 | | | | | | .005 | | | 1.830 | | | .165 | | | .340 | | |
| **TIV** | | | | .001 | | | <.001 | | | 4.903 | | | <.001 | | | .001 | | | | | | <.001 | | | 4.227 | | | .001* | | | .456 | | |
| **Age** | | | | -.001 | | | .007 | | | -.143 | | | .938 | | | -.009 | | | | | | .007 | | | -1.380 | | | .219 | | | .387 | | |
| **Model** | | | | N = 21, R² = .607, F = 8.7,  p = .001* | | | | | | | | | | | | | | | N = 24, R² = .603, F = 10.1,  p < .001* | | | | | | | | | | | | | | |
| **Left dorsal striatum** | | | | | | | | | | | | | | | | | | | | | | | | | | | | | | | | | |
|  | **E2 condition** | | | | | | | | | | | | | | | **PLAC condition** | | | | | | | | | | | | | | | **Z test** | | |
|  | | | | β | | | SE | | | t-stat | | | p*_FDR_* | | | β | | | | | | SE | | | t-stat | | | p*_FDR_* | | | p | | |
| **Reappraisal x E2Δ** | | | | .011 | | | .075 | | | .143 | | | .888 | | | -.017 | | | | | | .024 | | | -.717 | | | .723 | | | .724 | | |
| **TIV** | | | | .005 | | | .001 | | | 4.480 | | | .001* | | | .006 | | | | | | .001 | | | 7.210 | | | <.001* | | | .579 | | |
| **Age** | | | | -.018 | | | .040 | | | -.443 | | | .845 | | | -.013 | | | | | | .030 | | | -.418 | | | .817 | | | .922 | | |
| **Model** | | | | N = 21, R² = .582, F = 7.8,  p = .002* | | | | | | | | | | | | | | | N = 24, R² = .738, F = 18.4,  p < .001* | | | | | | | | | | | | | | |
| **Right dorsal striatum** | | | | | | | | | | | | | | | | | | | | | | | | | | | | | | | | | |
|  | **E2 condition** | | | | | | | | | | | | | | | **PLAC condition** | | | | | | | | | | | | | | | **Z test** | | |
|  | | | | β | | | SE | | | t-stat | | | p*_FDR_* | | | β | | | | | | SE | | | t-stat | | | p*_FDR_* | | | p | | |
| **Reappraisal x E2Δ** | | | | .031 | | | .081 | | | .386 | | | .845 | | | -.004 | | | | | | .026 | | | -.146 | | | .886 | | | .680 | | |
| **TIV** | | | | .005 | | | .001 | | | 4.482 | | | .001* | | | .006 | | | | | | .001 | | | 6.759 | | | <.001* | | | .727 | | |
| **Age** | | | | -.028 | | | .043 | | | -.653 | | | .845 | | | -.037 | | | | | | .033 | | | -1.109 | | | .561 | | | .870 | | |
| **Model** | | | | N = 21 R² = .594, F = 8.3,  p = .001* | | | | | | | | | | | | | | | N = 24, R² = .727, F = 17.5,  p < .001* | | | | | | | | | | | | | | |
| **Left caudoventral striatum** | | | | | | | | | | | | | | | | | | | | | | | | | | | | | | | | | |
|  | **E2 condition** | | | | | | | | | | | | | | | **PLAC condition** | | | | | | | | | | | | | | | **Z test** | | |
|  | | | | β | | | SE | | | t-stat | | | p*_FDR_* | | | β | | | | | | SE | | | t-stat | | | p*_FDR_* | | | p | | |
| **Reappraisal x E2Δ** | | | | -.002 | | | .005 | | | -.402 | | | .911 | | | <-.001 | | | | | | .002 | | | -.137 | | | .893 | | | .741 | | |
| **TIV** | | | | <.001 | | | <.001 | | | 4.489 | | | .002* | | | <.001 | | | | | | <.001 | | | 4.728 | | | <.001* | | | .671 | | |
| **Age** | | | | .001 | | | .003 | | | .225 | | | .911 | | | -.001 | | | | | | .002 | | | -.674 | | | .762 | | | .538 | | |
| **Model** | | | | N = 21, R² = .554, F = 7.0,  p = .003* | | | | | | | | | | | | | | | N = 24, R² = .558, F = 8.4,  p = .001* | | | | | | | | | | | | | | |
| **Right caudoventral striatum** | | | | | | | | | | | | | | | | | | | | | | | | | | | | | | | | | |
|  | **E2 condition** | | | | | | | | | | | | | | | **PLAC condition** | | | | | | | | | | | | | | | **Z test** | | |
|  | | | | β | | | SE | | | t-stat | | | p*_FDR_* | | | β | | | | | | SE | | | t-stat | | | p*_FDR_* | | | p | | |
| **Reappraisal x E2Δ** | | | | .006 | | | .005 | | | 1.249 | | | .457 | | | -.001 | | | | | | .002 | | | -.453 | | | .786 | | | .184 | | |
| **TIV** | | | | <.001 | | | <.001 | | | 2.777 | | | .038* | | | <.001 | | | | | | <.001 | | | 4.664 | | | <.001* | | | .340 | | |
| **Age** | | | | <-.001 | | | .003 | | | -.114 | | | .911 | | | -.002 | | | | | | .003 | | | -.935 | | | .722 | | | .581 | | |
| **Model** | | | | N = 21, R² = .407, F = 3.9,  p =.028* | | | | | | | | | | | | | | | N = 24, R² = .556, F = 8.3,  p < .001* | | | | | | | | | | | | | | |

Interaction E2 x rumination.

**Tab. S7 | Statistical parameters from robust mixed linear regression analysing** the relationship between emotion regulation strategy (trait reappraisal and rumination) by E2 increase and regional gray matter volume (GMV) under E2 and placebo (PLAC) conditions. Slope coefficients (β), standard errors (SE), t-statistics, and false discovery rate (FDR)-corrected p-values for the association between E2 increase and GMV across both drug conditions are reported. Additionally, p-values from Z-transformed slope comparisons (E2 vs. PLAC) are included to assess condition-specific differences in association strength. Significant associations (p < .05) are marked with an asterisk (*).

| **Left amygdala** | | | | | | | | | | | | | | | | | | | | | | | | | | | | | | | | | |
| --- | --- | --- | --- | --- | --- | --- | --- | --- | --- | --- | --- | --- | --- | --- | --- | --- | --- | --- | --- | --- | --- | --- | --- | --- | --- | --- | --- | --- | --- | --- | --- | --- | --- |
|  | | | **E2 condition** | | | | | | | | | | | | | | | **PLAC condition** | | | | | | | | | | | | | | | **Z test** |
|  | | | | | | β | | | SE | | | t-stat | | | p*_FDR_* | | | β | | | | | | SE | | | t-stat | | | p*_FDR_* | | | p |
| **Rumination x E2Δ** | | | | | | -.026 | | | .020 | | | -1.321 | | | .408 | | | -.005 | | | | | | .006 | | | -.877 | | | .782 | | | .305 |
| **TIV** | | | | | | .002 | | | <.001 | | | 5.297 | | | <.001* | | | .001 | | | | | | <.001 | | | 4.964 | | | <.001* | | | .566 |
| **Age** | | | | | | .005 | | | .011 | | | .470 | | | .644 | | | -.002 | | | | | | .011 | | | -.153 | | | .880 | | | .659 |
| **Model** | | | | | | N = 21, R² = .680, F = 12.0,  p = .001* | | | | | | | | | | | | | | | N = 24, R² = .497, F= 6.6,  p < .003* | | | | | | | | | | | | |
| **Right amygdala** | | | | | | | | | | | | | | | | | | | | | | | | | | | | | | | | | |
|  | | | **E2 condition** | | | | | | | | | | | | | | | **PLAC condition** | | | | | | | | | | | | | | | **Z test** |
|  | | | | | | β | | | SE | | | t-stat | | | p*_FDR_* | | | β | | | | | | SE | | | t-stat | | | p*_FDR_* | | | p |
| **Rumination x E2Δ** | | | | | | -.025 | | | .024 | | | -1.04 | | | .469 | | | -.004 | | | | | | .006 | | | -.600 | | | .833 | | | .390 |
| **TIV** | | | | | | .002 | | | <.001 | | | 5.335 | | | <.001* | | | .002 | | | | | | <.001 | | | 5.46 | | | <.001* | | | .464 |
| **Age** | | | | | | .010 | | | .013 | | | .762 | | | .548 | | | -.004 | | | | | | .011 | | | -.365 | | | .863 | | | .416 |
| **Model** | | | | | | N = 21, R² = .686, F = 12.3,  p < .001* | | | | | | | | | | | | | | | N = 24, R² = .626, F = 11.2,  p < .001* | | | | | | | | | | | | |
| **Left ACC** \| \| \| \| \| \| \| \| \| \| \|; \| **E2 condition** \| \| \| \| \| **PLAC condition** \| \| \| \| \| **Z test**; \| \| β \| SE \| t-stat \| p*_FDR_* \| β \| \| SE \| t-stat \| p*_FDR_* \| p; **Rumination x E2Δ** \| \| .080 \| .081 \| .990 \| .495 \| .006 \| \| .020 \| .269 \| .790 \| .371; **TIV** \| \| .006 \| .001 \| 4.722 \| .001* \| .006 \| \| .001 \| 6.177 \| <.001* \| .874; **Age** \| \| -.103 \| .044 \| -2.360 \| .061 \| -.071 \| \| .038 \| -1.883 \| .149 \| .587; **Model** \| \| N = 21, R² = .779, F = 20, p < .001* \| \| \| \| \| N = 24, R² = .733, F= 18.3, p < .001* \| \| \| \| | | | | | | | | | | | | | | | | | | | | | | | | | | | | | | | | | |
| **Right ACC** \| \| \| \| \| \| \| \| \| \| \|; \| **E2 condition** \| \| \| \| \| **PLAC condition** \| \| \| \| \| **Z test**; \| \| β \| SE \| t-stat \| p*_FDR_* \| β \| \| SE \| t-stat \| p*_FDR_* \| p; **Rumination x E2Δ** \| \| .006 \| .083 \| .072 \| .944 \| .009 \| \| .0171 \| .542 \| .713 \| .969; **TIV** \| \| .006 \| .001 \| 4.885 \| .001* \| .006 \| \| .001 \| 7.183 \| <.001* \| .853; **Age** \| \| -.038 \| .045 \| -.841 \| .495 \| -.032 \| \| .032 \| -.990 \| .501 \| .910; **Model** \| \| N = 21, R² = .721, F = 14.7, p < .001* \| \| \| \| \| N = 24, R² = .774, F = 22.8, p < .001* \| \| \| \| | | | | | | | | | | | | | | | | | | | | | | | | | | | | | | | | | |
| **Left perigenual ACC** | | | | | | | | | | | | | | | | | | | | | | | | | | | | | | | | | |
|  | | **E2 condition** | | | | | | | | | | | | | | | **PLAC condition** | | | | | | | | | | | | | | | **Z test** | |
|  | | | | | β | | | SE | | | t-stat | | | p*_FDR_* | | | β | | | | | | SE | | | t-stat | | | p*_FDR_* | | | p | |
| **Rumination x E2Δ** | | | | | .066 | | | .058 | | | 1.134 | | | .409 | | | .001 | | | | | | .015 | | | .066 | | | .949 | | | .281 | |
| **TIV** | | | | | .004 | | | .001 | | | 4.331 | | | .001* | | | .004 | | | | | | .001 | | | 5.572 | | | <.001* | | | .785 | |
| **Age** | | | | | -.079 | | | .031 | | | -2.505 | | | .045* | | | -.057 | | | | | | .029 | | | -1.964 | | | .127 | | | .605 | |
| **Model** | | | | | N = 21, R² = .765, F = 18.4,  p < .001* | | | | | | | | | | | | | | | N = 24, R² = .694, F = 15.1,  p < .001* | | | | | | | | | | | | | |
| **Right perigenual ACC** | | | | | | | | | | | | | | | | | | | | | | | | | | | | | | | | | |
|  | | **E2 condition** | | | | | | | | | | | | | | | **PLAC condition** | | | | | | | | | | | | | | | **Z test** | |
|  | | | | | β | | | SE | | | t-stat | | | p*_FDR_* | | | β | | | | | | SE | | | t-stat | | | *p_FDR_* | | | p | |
| **Rumination x E2Δ** | | | | | .008 | | | .060 | | | .131 | | | .897 | | | .008 | | | | | | .013 | | | .601 | | | .665 | | | .995 | |
| **TIV** | | | | | 004 | | | .001 | | | 4.406 | | | .001* | | | .004 | | | | | | .001 | | | 6.486 | | | <.001* | | | .909 | |
| **Age** | | | | | -.030 | | | .033 | | | -.925 | | | .442 | | | -.026 | | | | | | .023 | | | -1.119 | | | .414 | | | .922 | |
| **Model** | | | | | N = 21, R² = .686, F = 12.4,  p < .001* | | | | | | | | | | | | | | | N = 24, R² = .743, F = 19.3,  p < .001* | | | | | | | | | | | | | |
| **Left subgenual ACC** | | | | | | | | | | | | | | | | | | | | | | | | | | | | | | | | | |
|  | | **E2 condition** | | | | | | | | | | | | | | | **PLAC condition** | | | | | | | | | | | | | | | **Z test** | |
|  | | | | | β | | | SE | | | t-stat | | | p*_FDR_* | | | β | | | | | | SE | | | t-stat | | | p*_FDR_* | | | p | |
| **Rumination x E2Δ** | | | | | .022 | | | .028 | | | .793 | | | .658 | | | .005 | | | | | | .006 | | | .779 | | | .519 | | | .546 | |
| **TIV** | | | | | .002 | | | 4.287 | | | 3.480 | | | .009* | | | .001 | | | | | | <.001 | | | 4.790 | | | <.001* | | | .934 | |
| **Age** | | | | | -.020 | | | .015 | | | -1.333 | | | .400 | | | -.011 | | | | | | .012 | | | -.916 | | | .519 | | | .619 | |
| **Model** | | | | | N = 21, R² = .641, F = 10.1,  p < .001* | | | | | | | | | | | | | | | N = 24, R² = .629, F = 11.3,  p < .001* | | | | | | | | | | | | | |
| **Right subgenual ACC** | | | | | | | | | | | | | | | | | | | | | | | | | | | | | | | | | |
|  | | **E2 condition** | | | | | | | | | | | | | | | **PLAC condition** | | | | | | | | | | | | | | | **Z test** | |
|  | | | | | β | | | SE | | | t-stat | | | p*_FDR_* | | | β | | | | | | SE | | | t-stat | | | p*_FDR_* | | | p | |
| **Rumination x E2Δ** | | | | | -.007 | | | .022 | | | -.293 | | | .773 | | | .003 | | | | | | .005 | | | .656 | | | .519 | | | .675 | |
| **TIV** | | | | | .002 | | | <.001 | | | 4.870 | | | .001* | | | .001 | | | | | | <.001 | | | 6.222 | | | <.001* | | | .499 | |
| **Age** | | | | | -.007 | | | .012 | | | -.572 | | | .690 | | | -.008 | | | | | | .009 | | | -.932 | | | .519 | | | .940 | |
| **Model** | | | | | N = 21, R² = .700, F = 13.3,  p < .001* | | | | | | | | | | | | | | | N = 24, R² = .726, F = 17.7,  p < .001* | | | | | | | | | | | | | |
| **Left dorsal ACC** | | | | | | | | | | | | | | | | | | | | | | | | | | | | | | | | | |
|  | | **E2 condition** | | | | | | | | | | | | | | | **PLAC condition** | | | | | | | | | | | | | | | **Z test** | |
|  | | | | | β | | | SE | | | t-stat | | | p*_FDR_* | | | β | | | | | | SE | | | t-stat | | | p*_FDR_* | | | p | |
| **Rumination x E2Δ** | | | | | -.004 | | | .009 | | | -.465 | | | .860 | | | <.001 | | | | | | .002 | | | .136 | | | .894 | | | .629 | |
| **TIV** | | | | | .001 | | | <.001 | | | 3.827 | | | .008* | | | <.001 | | | | | | <.001 | | | 4.869 | | | .001* | | | .803 | |
| **Age** | | | | | -.006 | | | .005 | | | -1.330 | | | .402 | | | -.005 | | | | | | .004 | | | -1.478 | | | .310 | | | .897 | |
| **Model** | | | | | N = 21, R² = .630, F = 9.6,  p = .001* | | | | | | | | | | | | | | | N = 24, R² = 628, F = 11.3,  p < .001* | | | | | | | | | | | | | |
| **Right dorsal ACC** | | | | | | | | | | | | | | | | | | | | | | | | | | | | | | | | | |
|  | | **E2 condition** | | | | | | | | | | | | | | | **PLAC condition** | | | | | | | | | | | | | | | **Z test** | |
|  | | | | | β | | | SE | | | t-stat | | | p*_FDR_* | | | β | | | | | | SE | | | t-stat | | | p*_FDR_* | | | p | |
| **Rumination x E2Δ** | | | | | .002 | | | .011 | | | .179 | | | .860 | | | <-.001 | | | | | | .003 | | | -.179 | | | .894 | | | .827 | |
| **TIV** | | | | | <.001 | | | <.001 | | | 3.203 | | | .016* | | | <.001 | | | | | | <.001 | | | 3.781 | | | .004* | | | .931 | |
| **Age** | | | | | -.002 | | | .006 | | | -.300 | | | .860 | | | .001 | | | | | | .006 | | | .184 | | | .894 | | | .729 | |
| **Model** | | | | | N = 21, R² = .523, F = 6.2,  p = .005* | | | | | | | | | | | | | | | N = 24, R² = .446, F = 5.3,  p =.007* | | | | | | | | | | | | | |
| **Left total hippocampus** \| \| \| \| \| \| \| \| \| \| \|; \| **E2 condition** \| \| \| \| \| **PLAC condition** \| \| \| \| \| **Z test**; \| \| β \| SE \| t-stat \| p*_FDR_* \| β \| \| SE \| t-stat \| p*_FDR_* \| p; **Rumination x E2Δ** \| \| -.022 \| .060 \| -.361 \| .976 \| -.013 \| \| .013 \| -1.019 \| .641 \| .892; **TIV** \| \| .003 \| .002 \| 3.473 \| <.009* \| .003 \| \| .006 \| 4.797 \| <.001* \| .893; **Age** \| \| -.001 \| .032 \| -.031 \| .976 \| -.004 \| \| .024 \| -.148 \| .884 \| .949; **Model** \| \| N = 21, R² = .517, F = 6.08, p = .005* \| \| \| \| \| N = 24, R² = .549, F= 8.1, p = .001* \| \| \| \| | | | | | | | | | | | | | | | | | | | | | | | | | | | | | | | | | |
| **Right total hippocampus** \| \| \| \| \| \| \| \| \| \| \|; \| **E2 condition** \| \| \| \| \| **PLAC condition** \| \| \| \| \| **Z test**; \| \| β \| SE \| t-stat \| p*_FDR_* \| β \| \| SE \| t-stat \| p*_FDR_* \| p; **Rumination x E2Δ** \| \| -.005 \| .061 \| -.083 \| .976 \| -.011 \| \| .013 \| -.837 \| .875 \| .925; **TIV** \| \| .003 \| .001 \| 3.271 \| <.007* \| .003 \| \| .001 \| 4.644 \| <.001* \| .917; **Age** \| \| -.015 \| .033 \| -.436 \| .976 \| -.011 \| \| .025 \| -.458 \| .875 \| .937; **Model** \| \| N = 21, R² = .523, F = 6.2, p = .005* \| \| \| \| \| N = 24, R² = .543, F= 7.9, p = .001* \| \| \| \| | | | | | | | | | | | | | | | | | | | | | | | | | | | | | | | | | |
| **Left hippocampus** | | | | | | | | | | | | | | | | | | | | | | | | | | | | | | | | | |
|  | | **E2 condition** | | | | | | | | | | | | | | | **PLAC condition** | | | | | | | | | | | | | | | **Z test** | |
|  | | | | | β | | | SE | | | t-stat | | | p*_FDR_* | | | β | | | | | | SE | | | t-stat | | | p*_FDR_* | | | p | |
| **Rumination x E2Δ** | | | | | -.015 | | | .029 | | | -.516 | | | .961 | | | -.005 | | | | | | .007 | | | -.631 | | | .642 | | | .725 | |
| **TIV** | | | | | .002 | | | <.001 | | | 4.386 | | | .002* | | | .002 | | | | | | <.001 | | | 4.562 | | | .001* | | | .498 | |
| **Age** | | | | | .001 | | | .016 | | | .066 | | | .961 | | | -.009 | | | | | | .013 | | | -.672 | | | .642 | | | .629 | |
| **Model** | | | | | N = 21, R² = .625, F = 9.5,  p = .001* | | | | | | | | | | | | | | | N = 24, R² = .546, F = 8.0,  p = .001* | | | | | | | | | | | | | |
| **Right hippocampus** | | | | | | | | | | | | | | | | | | | | | | | | | | | | | | | | | |
|  | | **E2 condition** | | | | | | | | | | | | | | | **PLAC condition** | | | | | | | | | | | | | | | **Z test** | |
|  | | | | | β | | | SE | | | t-stat | | | p*_FDR_* | | | β | | | | | | SE | | | t-stat | | | p*_FDR_* | | | p | |
| **Rumination x E2Δ** | | | | | -.003 | | | .037 | | | -.075 | | | .961 | | | -.006 | | | | | | .009 | | | -.726 | | | .642 | | | .929 | |
| **TIV** | | | | | .002 | | | .001 | | | 3.273 | | | .014* | | | .002 | | | | | | <.001 | | | 3.983 | | | .002* | | | .762 | |
| **Age** | | | | | .001 | | | .020 | | | .050 | | | .961 | | | <-.001 | | | | | | .016 | | | -.027 | | | .979 | | | .956 | |
| **Model** | | | | | N = 21, R² = .502, F = 5.7,  p = .007* | | | | | | | | | | | | | | | N = 24, R² = .457, F = 5.6,  p = .006* | | | | | | | | | | | | | |
| **Left para-hippocampus** | | | | | | | | | | | | | | | | | | | | | | | | | | | | | | | | | |
|  | | **E2 condition** | | | | | | | | | | | | | | | **PLAC condition** | | | | | | | | | | | | | | | **Z test** | |
|  | | | | | β | | | SE | | | t-stat | | | p*_FDR_* | | | β | | | | | | SE | | | t-stat | | | p*_FDR_* | | | p | |
| **Rumination x E2Δ** | | | | | -.009 | | | .035 | | | -.250 | | | .993 | | | -.009 | | | | | | .007 | | | -1.297 | | | .419 | | | .991 | |
| **TIV** | | | | | .001 | | | .001 | | | 2.298 | | | .104* | | | .002 | | | | | | <.001 | | | 4.395 | | | <.001* | | | .665 | |
| **Age** | | | | | <-.001 | | | .019 | | | -.009 | | | .993 | | | .011 | | | | | | .013 | | | .853 | | | .490 | | | .620 | |
| **Model** | | | | | N = 21, R² = .318, F = 2.6,  p = .083 | | | | | | | | | | | | | | | N = 24, R² = .493, F = 6.5,  p = .003* | | | | | | | | | | | | | |
| **Right para-hippocampus** | | | | | | | | | | | | | | | | | | | | | | | | | | | | | | | | | |
|  | | **E2 condition** | | | | | | | | | | | | | | | **PLAC condition** | | | | | | | | | | | | | | | **Z test** | |
|  | | | | | β | | | SE | | | t-stat | | | p*_FDR_* | | | β | | | | | | SE | | | t-stat | | | p*_FDR_* | | | p | |
| **Rumination x E2Δ** | | | | | .003 | | | .030 | | | .099 | | | .993 | | | -.005 | | | | | | .007 | | | -.828 | | | .490 | | | .785 | |
| **TIV** | | | | | .001 | | | <.001 | | | 2.592 | | | .104 | | | .001 | | | | | | <.001 | | | 4.383 | | | <.001* | | | .732 | |
| **Age** | | | | | -.015 | | | .016 | | | -.928 | | | .733 | | | -.009 | | | | | | .012 | | | -.703 | | | .490 | | | .749 | |
| **Model** | | | | | N = 21, R² = .460, F = 4.8,  p = .013* | | | | | | | | | | | | | | | N = 24, R² = .521, F = 7.3,  p = .002* | | | | | | | | | | | | | |
| (**Left striatum** \| \| \| \| \| \| \| \| \| \| \|; \| **E2 condition** \| \| \| \| \| **PLAC condition** \| \| \| \| \| **Z test**; \| \| β \| SE \| t-stat \| p*_FDR_* \| β \| \| SE \| t-stat \| p*_FDR_* \| p; **Rumination x E2Δ** \| \| <.001 \| .094 \| .005 \| .996 \| -.008 \| \| .019 \| -.413 \| .765 \| .931; **TIV** \| \| .006 \| .001 \| 4.105 \| .003* \| .007 \| \| .001 \| 7.059 \| <.001* \| .721; **Age** \| \| -.009 \| .051 \| -.181 \| .996 \| -.013 \| \| .036 \| -.351 \| .765 \| .958; **Model** \| \| N = 21, R² = .629, F = 9.6, p = .001* \| \| \| \| \| N = 24, R² = .744, F = 19.4, p < .001* \| \| \| \|) | | | | | | | | | | | | | | | | | | | | | | | | | | | | | | | | | |
| **Right striatum** \| \| \| \| \| \| \| \| \| \| \|; \| **E2 condition** \| \| \| \| \| **PLAC condition** \| \| \| \| \| **Z test**; \| \| β \| SE \| t-stat \| p*_FDR_* \| β \| \| SE \| t-stat \| p*_FDR_* \| p; **Rumination x E2Δ** \| \| -.016 \| .105 \| -.150 \| .996 \| -.006 \| \| .021 \| -.304 \| .765 \| .929; **TIV** \| \| .006 \| .002 \| 3.930 \| .003* \| .006 \| \| .001 \| 6.327 \| <.001* \| .986; **Age** \| \| -.028 \| .057 \| -.490 \| .996 \| -.053 \| \| .038 \| -1.372 \| .370 \| .718; **Model** \| \| N = 21, R² = .709, F = 8.8, p = .001* \| \| \| \| \| N = 24, R² = .721, F = 17.3, p < .001* \| \| \| \| | | | | | | | | | | | | | | | | | | | | | | | | | | | | | | | | | |
| **Left ventral striatum** | | | | | | | | | | | | | | | | | | | | | | | | | | | | | | | | | |
|  | **E2 condition** | | | | | | | | | | | | | | | **PLAC condition** | | | | | | | | | | | | | | | **Z test** | | |
|  | | | | β | | | SE | | | t-stat | | | p*_FDR_* | | | β | | | | | | SE | | | t-stat | | | p*_FDR_* | | | p | | |
| **Rumination x E2Δ** | | | | .007 | | | .016 | | | .455 | | | .751 | | | .005 | | | | | | .003 | | | 1.603 | | | .213 | | | .890 | | |
| **TIV** | | | | .001 | | | <.001 | | | 2.507 | | | .066 | | | .001 | | | | | | <.001 | | | 4.631 | | | .001* | | | .761 | | |
| **Age** | | | | -.008 | | | .009 | | | -.955 | | | .706 | | | -.004 | | | | | | .006 | | | -.632 | | | .535 | | | .662 | | |
| **Model** | | | | N = 21, R² = .472, F = 5.1,  p = .011* | | | | | | | | | | | | | | | N = 24, R² = .647, F = 12.2,  p < .001* | | | | | | | | | | | | | | |
| **Right ventral striatum** | | | | | | | | | | | | | | | | | | | | | | | | | | | | | | | | | |
|  | **E2 condition** | | | | | | | | | | | | | | | **PLAC condition** | | | | | | | | | | | | | | | **Z test** | | |
|  | | | | β | | | SE | | | t-stat | | | p*_FDR_* | | | β | | | | | | SE | | | t-stat | | | p*_FDR_* | | | p | | |
| **Rumination x E2Δ** | | | | -.005 | | | .014 | | | -.359 | | | .751 | | | .006 | | | | | | .004 | | | 1.530 | | | .213 | | | .461 | | |
| **TIV** | | | | .001 | | | <.001 | | | 4.424 | | | .002* | | | .001 | | | | | | <.001 | | | 3.810 | | | .003* | | | .338 | | |
| **Age** | | | | .003 | | | .008 | | | .322 | | | .751 | | | -.009 | | | | | | .007 | | | -1.281 | | | .258 | | | .271 | | |
| **Model** | | | | N = 21, R² = .633, F = 9.6,  p = .001* | | | | | | | | | | | | | | | N = 24, R² = .595, F = 9.8,  p < .001* | | | | | | | | | | | | | | |
| **Left dorsal striatum** | | | | | | | | | | | | | | | | | | | | | | | | | | | | | | | | | |
|  | **E2 condition** | | | | | | | | | | | | | | | **PLAC condition** | | | | | | | | | | | | | | | **Z test** | | |
|  | | | | β | | | SE | | | t-stat | | | p*_FDR_* | | | β | | | | | | SE | | | t-stat | | | p*_FDR_* | | | p | | |
| **Rumination x E2Δ** | | | | .015 | | | .085 | | | .176 | | | .956 | | | -.012 | | | | | | .016 | | | -.797 | | | .585 | | | .759 | | |
| **TIV** | | | | .005 | | | .001 | | | 3.408 | | | .010* | | | .006 | | | | | | .001 | | | 6.896 | | | <.001* | | | .495 | | |
| **Age** | | | | -.032 | | | .046 | | | -.705 | | | .824 | | | -.014 | | | | | | .031 | | | -.449 | | | .658 | | | .736 | | |
| **Model** | | | | N = 21, R² = .572, F = 7.5,  p = .002* | | | | | | | | | | | | | | | N = 24, R² = .732, F = 17.9,  p < .001* | | | | | | | | | | | | | | |
| **Right dorsal striatum** | | | | | | | | | | | | | | | | | | | | | | | | | | | | | | | | | |
|  | **E2 condition** | | | | | | | | | | | | | | | **PLAC condition** | | | | | | | | | | | | | | | **Z test** | | |
|  | | | | β | | | SE | | | t-stat | | | p*_FDR_* | | | β | | | | | | SE | | | t-stat | | | p*_FDR_* | | | p | | |
| **Rumination x E2Δ** | | | | -.005 | | | .091 | | | -.056 | | | .956 | | | -.014 | | | | | | .017 | | | -.808 | | | .585 | | | .925 | | |
| **TIV** | | | | .005 | | | .001 | | | 3.677 | | | .010* | | | .005 | | | | | | .001 | | | 6.098 | | | <.001* | | | .957 | | |
| **Age** | | | | -.030 | | | .049 | | | -.611 | | | .824 | | | -.056 | | | | | | .032 | | | -1.758 | | | .188 | | | .659 | | |
| **Model** | | | | N = 21, R² = .588, F = 8.1,  p = .002* | | | | | | | | | | | | | | | N =24, R² = .707, F = 15.8,  p < .001* | | | | | | | | | | | | | | |
| **Left caudoventral striatum** | | | | | | | | | | | | | | | | | | | | | | | | | | | | | | | | | |
|  | **E2 condition** | | | | | | | | | | | | | | | **PLAC condition** | | | | | | | | | | | | | | | **Z test** | | |
|  | | | | β | | | SE | | | t-stat | | | p*_FDR_* | | | β | | | | | | SE | | | t-stat | | | p*_FDR_* | | | p | | |
| **Rumination x E2Δ** | | | | -.003 | | | .005 | | | -.524 | | | .847 | | | -.001 | | | | | | .001 | | | -1.190 | | | .372 | | | .789 | | |
| **TIV** | | | | <.001 | | | <.001 | | | 3.672 | | | .011* | | | <.001 | | | | | | <.001 | | | 5.145 | | | <.001* | | | .838 | | |
| **Age** | | | | <.001 | | | .003 | | | .173 | | | .865 | | | -.002 | | | | | | .002 | | | -.788 | | | .440 | | | .546 | | |
| **Model** | | | | N = 21, R² = .531, F = 6.4,  p = .004* | | | | | | | | | | | | | | | N = 24, R² = .595, F = 9.8,  p < .001* | | | | | | | | | | | | | | |
| **Right caudoventral striatum** | | | | | | | | | | | | | | | | | | | | | | | | | | | | | | | | | |
|  | **E2 condition** | | | | | | | | | | | | | | | **PLAC condition** | | | | | | | | | | | | | | | **Z test** | | |
|  | | | | β | | | SE | | | t-stat | | | p*_FDR_* | | | β | | | | | | SE | | | t-stat | | | p*_FDR_* | | | p | | |
| **Rumination x E2Δ** | | | | -.007 | | | .006 | | | -1.283 | | | .433 | | | -.002 | | | | | | .001 | | | -1.331 | | | .372 | | | .336 | | |
| **TIV** | | | | <.001 | | | <.001 | | | 3.182 | | | .016* | | | <.001 | | | | | | <.001 | | | 5.040 | | | <.001 | | | .750 | | |
| **Age** | | | | .001 | | | .004 | | | .384 | | | .847 | | | -.003 | | | | | | .002 | | | -1.033 | | | .377 | | | .349 | | |
| **Model** | | | | N = 21, R² = .412, F = 4.0,  p =.026* | | | | | | | | | | | | | | | N = 24, R² = .591, F = 9.6,  p < .001* | | | | | | | | | | | | | | |
